# Rare predicted loss-of-function variants of type I IFN immunity genes are associated with life-threatening COVID-19

**DOI:** 10.1101/2022.10.22.22281221

**Authors:** Daniela Matuozzo, Estelle Talouarn, Astrid Marchal, Jeremy Manry, Yoann Seeleuthner, Yu Zhang, Alexandre Bolze, Matthieu Chaldebas, Baptiste Milisavljevic, Peng Zhang, Adrian Gervais, Paul Bastard, Takaki Asano, Lucy Bizien, Federica Barzaghi, Hassan Abolhassani, Ahmad Abou Tayoun, Alessandro Aiuti, Ilad Alavi Darazam, Luis M. Allende, Rebeca Alonso-Arias, Andrés Augusto Arias, Gokhan Aytekin, Peter Bergman, Simone Bondesan, Yenan T. Bryceson, Ingrid G. Bustos, Oscar Cabrera-Marante, Sheila Carcel, Paola Carrera, Giorgio Casari, Khalil Chaïbi, Roger Colobran, Antonio Condino-Neto, Laura E. Covill, Loubna El Zein, Carlos Flores, Peter K. Gregersen, Marta Gut, Filomeen Haerynck, Rabih Halwani, Selda Hancerli, Lennart Hammarström, Nevin Hatipoğlu, Adem Karbuz, Sevgi Keles, Christèle Kyheng, Rafael Leon-Lopez, Jose Luis Franco, Davood Mansouri, Javier Martinez-Picado, Ozge Metin Akcan, Isabelle Migeotte, Pierre-Emmanuel Morange, Guillaume Morelle, Andrea Martin-Nalda, Giuseppe Novelli, Antonio Novelli, Tayfun Ozcelik, Figen Palabiyik, Qiang Pan-Hammarström, Rebeca Pérez de Diego, Laura Planas-Serra, Daniel E. Pleguezuelo, Carolina Prando, Aurora Pujol, Luis Felipe Reyes, Jacques G. Rivière, Carlos Rodriguez-Gallego, Julian Rojas, Patrizia Rovere-Querini, Agatha Schlüter, Mohammad Shahrooei, Ali Sobh, Pere Soler-Palacin, Yacine Tandjaoui-Lambiotte, Imran Tipu, Cristina Tresoldi, Jesus Troya, Diederik van de Beek, Mayana Zatz, Pawel Zawadzki, Saleh Zaid Al-Muhsen, Hagit Baris-Feldman, Manish J. Butte, Stefan N. Constantinescu, Megan A. Cooper, Clifton L. Dalgard, Jacques Fellay, James R. Heath, Yu-Lung Lau, Richard P. Lifton, Tom Maniatis, Trine H. Mogensen, Horst von Bernuth, Alban Lermine, Michel Vidaud, Anne Boland, Jean-François Deleuze, Robert Nussbaum, Amanda Kahn-Kirby, France Mentre, Sarah Tubiana, Guy Gorochov, Florence Tubach, Pierre Hausfater, COVID Human Genetic Effort, COVIDeF Study Group, French COVID Cohort Study Group, CoV-Contact Cohort, COVID-STORM Clinicians, COVID Clinicians, Orchestra Working Group, Amsterdam UMC Covid-19 Biobank, NIAID-USUHS COVID Study Group, Isabelle Meyts, Shen-Ying Zhang, Anne Puel, Luigi D. Notarangelo, Stephanie Boisson-Dupuis, Helen C. Su, Bertrand Boisson, Emmanuelle Jouanguy, Jean-Laurent Casanova, Qian Zhang, Laurent Abel, Aurélie Cobat

## Abstract

**Background:** We previously reported inborn errors of TLR3- and TLR7-dependent type I interferon (IFN) immunity in 1-5% of unvaccinated patients with life-threatening COVID-19, and auto-antibodies against type I IFN in another 15-20% of cases.

**Methods:** We report here a genome-wide rare variant burden association analysis in 3,269 unvaccinated patients with life-threatening COVID-19 (1,301 previously reported and 1,968 new patients), and 1,373 unvaccinated SARS-CoV-2-infected individuals without pneumonia. A quarter of the patients tested had antibodies against type I IFN (234 of 928) and were excluded from the analysis.

**Results:** No gene reached genome-wide significance. Under a recessive model, the most significant gene with at-risk variants was *TLR7*, with an OR of 27.68 (95%CI:1.5-528.7, *P=*1.1×10^−4^), in analyses restricted to biochemically loss-of-function (bLOF) variants. We replicated the enrichment in rare predicted LOF (pLOF) variants at 13 influenza susceptibility loci involved in TLR3-dependent type I IFN immunity (OR=3.70 [95%CI:1.3-8.2], *P=*2.1×10^−4^). Adding the recently reported *TYK2* COVID-19 locus strengthened this enrichment, particularly under a recessive model (OR=19.65 [95%CI:2.1-2635.4]; *P=*3.4×10^−3^). When these 14 loci and *TLR7* were considered, all individuals hemizygous (*n*=20) or homozygous (*n*=5) for pLOF or bLOF variants were patients (OR=39.19 [95%CI:5.2-5037.0], *P*=4.7×10^−7^), who also showed an enrichment in heterozygous variants (OR=2.36 [95%CI:1.0-5.9], *P*=0.02). Finally, the patients with pLOF or bLOF variants at these 15 loci were significantly younger (mean age [SD]=43.3 [20.3] years) than the other patients (56.0 [17.3] years; *P=*1.68×10^−5^).

**Conclusions:** Rare variants of TLR3- and TLR7-dependent type I IFN immunity genes can underlie life-threatening COVID-19, particularly with recessive inheritance, in patients under 60 years old.

## Background

Clinical variability is high in unvaccinated individuals infected with severe acute respiratory syndrome coronavirus 2 (SARS-CoV-2), ranging from silent infection to lethal disease. In ∼ 3% of cases, infection leads to critical COVID-19 pneumonia, requiring high-flow oxygen (O_2_> 6 L/min), mechanical ventilation (non-invasive or by intubation), or extracorporeal membrane oxygenation (ECMO) [1]. Advanced age is by far the strongest predictor of COVID-19 severity, with the risk of death doubling every five years of age from childhood onward [2,3]. Men are also at greater risk of death than women [3–5]. Genome-wide (GW) association studies have identified several common loci associated with COVID-19 severity, the most significant being a region on chromosome 3p21.31 that was introduced by archaic introgression from Neanderthals [6–10]. The risk haplotype encompasses six genes and confers an estimated OR per copy of between 1.6 and 2.1, with higher values for individuals under 60 years old [7,11]. Twenty-four GW regions have been shown to be significantly associated with critical COVID-19 [10–12]. Four of these regions encompass genes involved in type I IFN immunity. The first, on chr12q24.13, containing protective variants, is also a Neanderthal haplotype and includes the *OAS1, OAS2*, and *OAS3* cluster, which are interferon stimulated genes (ISGs) required for the activation of antiviral RNaseL [13]. The second, a region on chr21q22.1, includes *IFNAR2*. The third, a region on chr19p13.2, includes *TYK2*. The fourth, a region on chr9p21, includes *IFNA10*. However, common variants have a modest effect size and explain only a very small fraction of the clinical variability [6,8]. This prompted us to search for rare variants conferring a stronger predisposition to life-threatening COVID-19.

Through a candidate approach focusing on influenza susceptibility genes, the COVID Human Genetics Effort (CHGE, www.covidhge.com) provided proof-of-concept that autosomal inborn errors of TLR3-dependent and -independent type I interferon (IFN) immunity, including autosomal recessive (AR) deficiencies of IFNAR1 or IRF7, can underlie critical COVID-19 [14]. Other children with AR IFNAR1, IFNAR2, TBK1, or STAT2 deficiency were subsequently reported, as well as children with AR TYK2 deficiency [15–19] (Figure 1). Some other groups were unable to replicate these findings, but the variants were not tested biochemically and it is unclear whether recessive defects were considered [11,20–22]. There may also be other reasons for their findings [1,23], the most important being the age distribution of the case cohorts. The other case cohorts were much older than ours (mean age of 66 vs. 52 years) and we found that inborn errors of immunity (IEI) were more frequent in patients under 60 years old. Consistently, we recently reported that ∼10% of children with moderate, severe, or critical COVID-19 pneumonia had recessive inborn errors of type I IFN immunity [18]. Moreover, older patients are more likely to carry pre-existing autoantibodies (auto-Abs) neutralizing type I IFN, which are found in about 15% of critical cases and up to 21% of patients over the age of 80 years [24,25]. The presence of such auto-Abs has been replicated by at least 26 studies worldwide [26,27], and we also recently showed that autoimmunity to type I IFNs is a strong common predictor of COVID-19 death in unvaccinated individuals, providing further evidence for the role of type I IFN immunity in life-threatening COVID-19.

**Figure 1.**
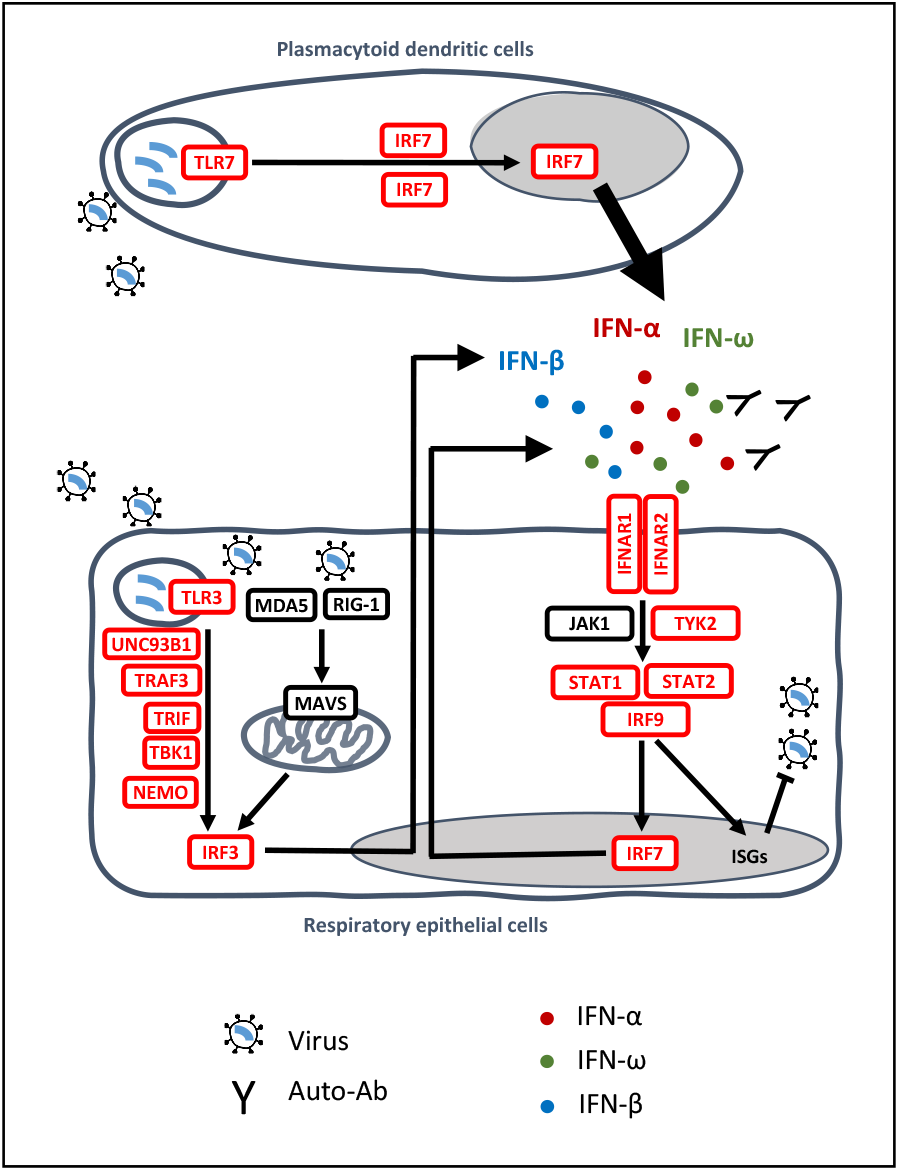
Type I IFN immunity genes associated with life-threatening COVID-19. Inborn errors of type I IFN immunity and autoantibodies neutralizing type I IFNs (α, β, ω) underlie life-threatening COVID-19 pneumonia by interfering with type I IFN immunity in respiratory epithelial cells (RECs) and blood plasmacytoid dendritic cells (pDCs). SARS-CoV-2 infection can induce type I IFN production in a TLR3-dependent manner in tissue-resident RECs (which express TLR3 but not TLR7) and in a TLR7-dependent manner in circulating pDCs (which express TLR7 but not TLR3). IRF7 is constitutively expressed in pDCs, at higher levels than in other cell types, whereas it is mostly induced by viral infection in RECs. Reported in red are the 13 genes (*IFNAR1, IFNAR2, IRF3, IRF7, IRF9, IKBKG, STAT1, STAT2, TBK1, TICAM1, TLR3, TRAF3* and *UNC93B1*) investigated in a previous study [14]; *TYK2* and *TLR7* were subsequently shown to underlie severe COVID-19 [18,28].

Using an unbiased X-wide gene burden test, we also identified X-linked recessive (XR) TLR7 deficiency in 17 male patients aged 7-71 years with critical COVID-19 pneumonia, accounting for ∼1% of cases in men (Figure 1) [28]. Moreover, six of the 11 *TLR7* variants previously reported in patients from other studies were deleterious (carried by nine of 16 patients) [29–34], whereas the *TLR7* variants in other studies were not disclosed [20,21]. TLR3 senses viral dsRNA in respiratory epithelial cells, whereas TLR7 senses ssRNA in plasmacytoid dendritic cells [1]. Both pathways induce the production of type I IFNs. *TLR7* gain-of-function variants were recently shown to be associated with human systemic lupus erythematosus [35], providing an example of mirror genetic effects between infectious and inflammatory/autoimmune diseases [36]. Collectively, these findings suggest that type I IFNs are essential for protective immunity to SARS-CoV-2 in the respiratory tract, with insufficient type I IFN activity accounting for up to 15-20% of cases of life-threatening COVID-19. Despite this high proportion, the determinants of critical COVID-19 pneumonia remain to be identified in ∼80% of cases. Here, we tested the hypotheses that other IEI may underlie critical COVID-19 pneumonia in at least some patients, and that our initial findings could be replicated in a new cohort. With the CHGE, we performed a GW gene-based rare variant association analysis. This analysis was performed in both previously investigated patients who had not been screened at the GW level [14,18,28], and in newly recruited patients.

## Materials and Methods

### Cohort

Since the beginning of the pandemic, we have enrolled more than 9,000 individuals with SARS-CoV2 infection and broad clinical manifestations from all over the world through the COVID Human Genetic Effort (CHGE). In this study we focused on patients with life-threatening COVID-19 and asymptomatic/mild infection. Life-threatening COVID-19 cases are defined as patients with pneumonia who developed critical disease, whether pulmonary with high-flow oxygen (>6 liter/min) or mechanical ventilation [continuous positive airway pressure (CPAP), bilevel positive airway pressure (BIPAP), and intubation], septic shock, or any other type of organ damage requiring intensive care unit admission (N=3503). We screened for the presence of autoantibodies (auto-Abs) against type I IFNs all patients for whom plasma was available (N=928) as previously described [24,25]. We excluded from the present analysis 234 patients who tested positive for auto-Abs as they already have a major risk factor for developing critical COVID-19 [27]. Controls are defined as individuals infected with SARS-CoV-2 who remained asymptomatic or pauci-symptomatic, with the presence of mild, self-healing, ambulatory disease (N=1373). Presence of infection has been assessed based on a positive PCR test and/or serological test and/or the presence of typical symptoms such as anosmia or agueusia after exposure to a confirmed COVID-19 case. Cases and controls were whole-exome (N= 2003 cases and 866 controls) or whole-genome (N=1266 cases and 507 controls) sequenced and high quality variants were obtained from the sequencing data as detailed in the Supplementary Methods.

### Population stratification

Principal component analysis (PCA) was performed with PLINK v1.9 software [37] on a pruned subset of ∼14,600 SNPs in linkage equilibrium (maximum r2 value of linkage disequilibrium 0.4 between pairs of SNPs) with minor allele frequency (MAF) > 1%, call rate > 99% and P value for departure from Hardy-Weinberg equilibrium > 10^−5^ as previously performed [38]. Ethnic origin was inferred from the PCA as previously described [38].

### Variant selection

For each gene, we considered several sets of candidate coding variants, defined according to (i) the functional annotation: predicted loss-of-function (pLOF) variants only (including stop gain/lost, start lost, frameshift, or splice variants), or pLOF with missense and inframe variants (MISSLOF); (ii) the Gnomad v2.1 allele frequency: variants with a Gnomad allele frequency below 1%, 0,1%, or 0,01%; and (iii) the CADD score for missense and inframe variants: missense and inframe variants with CADD score ≥ MSC for the corresponding gene or all the variants regardless of the CADD score.

### Rare variants burden analysis

We performed a genome-wide gene-based rare variants burden analysis. For each gene, the genotypic information at the candidate rare variants was summarized into a genetic score defined according to three genetic models: (i) co-dominant: samples are coded 2 if they carry at least one homozygous variant, 1 if they carry at least one heterozygous variant and 0 otherwise; (2) heterozygous: samples are coded 1 if they carry at least one heterozygous variant and 0 otherwise; and (3) recessive: samples are coded 1 if they carry at least one homozygous variant and 0 otherwise. For the X chromosome, hemizygous males are considered as homozygous females. Association between the genetic score for each gene and the disease status was tested by means of logistic regression-based likelihood ratio test (LRT) using EPACTS (Efficient and Parallelizable Association Container Toolbox) (http://genome.sph.umich.edu/wiki/EPACTS) for the genome-wide burden analysis or R 3.6.0 (https://cran.r-project.org/) for the candidate type I IFN related pathway. Firth’s bias correction, using the fast.logistf.fit function of EPACTS or the logistf function of the R logistf package, was applied if the *P value* of the LRT was below 0.05. Analyses were adjusted on sex, age (in years) and five first PCs of the PCA In Firth’s regression, a penalty term is placed on the standard maximum likelihood function used to estimate parameters of a logistic regression model when there are rare events or when complete separation exists [39]. With no covariates, this corresponds to adding 0.5 in every cell of a 2 by 2 table of allele counts versus case-control status. For a given gene and variant set, the burden test was not performed if the number of carriers across all samples was lower than 3.

We used three analysis strategies: 1) joint analysis of all samples; 2) trans-ethnic meta-analysis: the analysis was stratified according to 7 inferred ancestry subgroups (African, North African, European, admixed American, Middle Eastern, South Asian, East Asian). For each subgroup, an ethnic specific PCA was computed and used in the logistic regression model; and 3) trans-pipeline meta-analysis to account for the heterogeneity due to the type of sequencing data: the analysis was stratified according to the type of data shared (FASTQ vs VCF). Subgroups *P* values were further meta-analyzed accounting for the direction of effect and sample size using METAL [40].

### Multiple testing correction

For each gene, up to 18 burden tests were performed. Because these tests were not independent, we assessed the effective number of burden tests Meff by a method adapted from Patin et al. [41], based on the approach of Li and Ji [42]. This approach makes use of the variance of the eigenvalues of the observed statistics correlation matrix to estimate Meff. The Bonferroni corrected threshold was then defined as 0.05/Meff.

### Odds ratio (OR) equality for homozygous/hemizygous versus heterozygous carriers of pLOF variants at type I IFN genes

We tested whether the odd of critical COVID-19 for carriers versus non-carriers of pLOF variants at the type I IFN differed according to the zygosity status (homozygous/hemizygous vs heterozygous). In the full sample, we compared using LRT a full Firth bias corrected logistic regression model including two different parameters for carriers of pLOF according to the zygosity status (alternative hypothesis) with a Firth bias corrected logistic regression model including only one parameter for carriers of pLOF regardless of the zygosity status (null hypothesis). Analysis was performed with the R logistf package.

## Results

### Cohort description

Through the CHGE, we collected whole-exome sequencing (WES) or whole-genome sequencing (WGS) data for 3,503 patients with life-threatening COVID-19 pneumonia (hereafter referred to as “patients”; see Supplemental Methods) and 1,373 individuals with mild or asymptomatic infection, i.e. without pneumonia (hereafter referred to as “controls”). In total, 928 of the 3,503 patients were screened for the presence of auto-Abs to type I IFN [24,25] (Supplemental methods) and the 234 patients who tested positive were excluded from this analysis as they already have a major risk factor to develop critical COVID-19 [27]. In total, 1,301 of the 3,269 remaining patients had been included in previous studies restricted to a short list of 18 candidate genes [14,18] or to the X chromosome [28], and 1,968 had not been studied before. The mean age (SD) of the patients was 55.7 (17.4) years, with a male-to-female ratio of 2.4 (Table 1). The controls were significantly younger than the patients (*P* < 0.0001), with a mean age (SD) of 43.8 years (20.1 years) and were more likely to be female (*P* < 0.0001; male-to-female ratio = 0.7). The patients and controls were of various ethnic origins, mostly of European and Middle Eastern ancestry, according to principal component analysis (PCA) (Figure 2). Raw sequencing data were either centralized in the HGID laboratory and processed with the HGID pipeline (2,492 cases and 870 controls) or processed separately by each sequencing hub (777 cases and 503 controls; See Supplemental Methods). A joint analysis was performed first on the combined sample of 3,269 patients and 1,373 controls. Given the heterogeneity of the cohort due to different ancestries and processing pipelines, we also performed a trans-ethnic and a trans-pipeline meta-analysis; only results consistent across the three analyses are reported here (See Supplemental Methods).

**Table 1.** Baseline characteristics of study participants.

|  | Life-threatening COVID-19 | Infected controls | *P* value<sup>a</sup> |
| --- | --- | --- | --- |
|  | *n* = 3269 | *n* = 1373 |  |
| Sex, no. (%) |  |  |  |
| Male | 2314 (70.8%) | 542 (39.5%) | <0.0001 |
| Female | 955 (29.2%) | 831 (60.5%) |  |
| Age (years) |  |  |  |
| Mean (SD) | 55.74 (17.40) | 43.83 (20.14) | <0.0001 |
| Median (range) | 57 (0.08-99) | 43 (0.08-105) |  |
| Processing pipeline, no. (%) |  |  |  |
| HGID laboratory<sup>b</sup> | 2492 (76.2%) | 870 (63.4%) | <0.0001 |
| Other | 777 (24.8%) | 503 (36.6%) |  |
| Ancestry, no. (%) |  |  |  |
| European | 1374 (42.0%) | 960 (69.9%) | <0.0001 |
| Middle Eastern | 483 (14.8%) | 158 (11.5%) |  |
| Admixed American | 466 (14.3%) | 109 (7.9%) |  |
| North African | 300 (9.2%) | 24 (1.7%) |  |
| South Asian | 279 (8.5%) | 36 (2.6%) |  |
| Sub-Saharan African | 234 (7.1%) | 43 (3.1%) |  |
| East Asian | 133 (4.1%) | 43 (3.1%) |  |

<sup>a</sup> Chi-squared tests were used to compare proportions, and *t* tests were used to compare the mean ages.
<sup>b</sup>HGID: Human Genetics of Infectious Diseases

**Figure 2.**
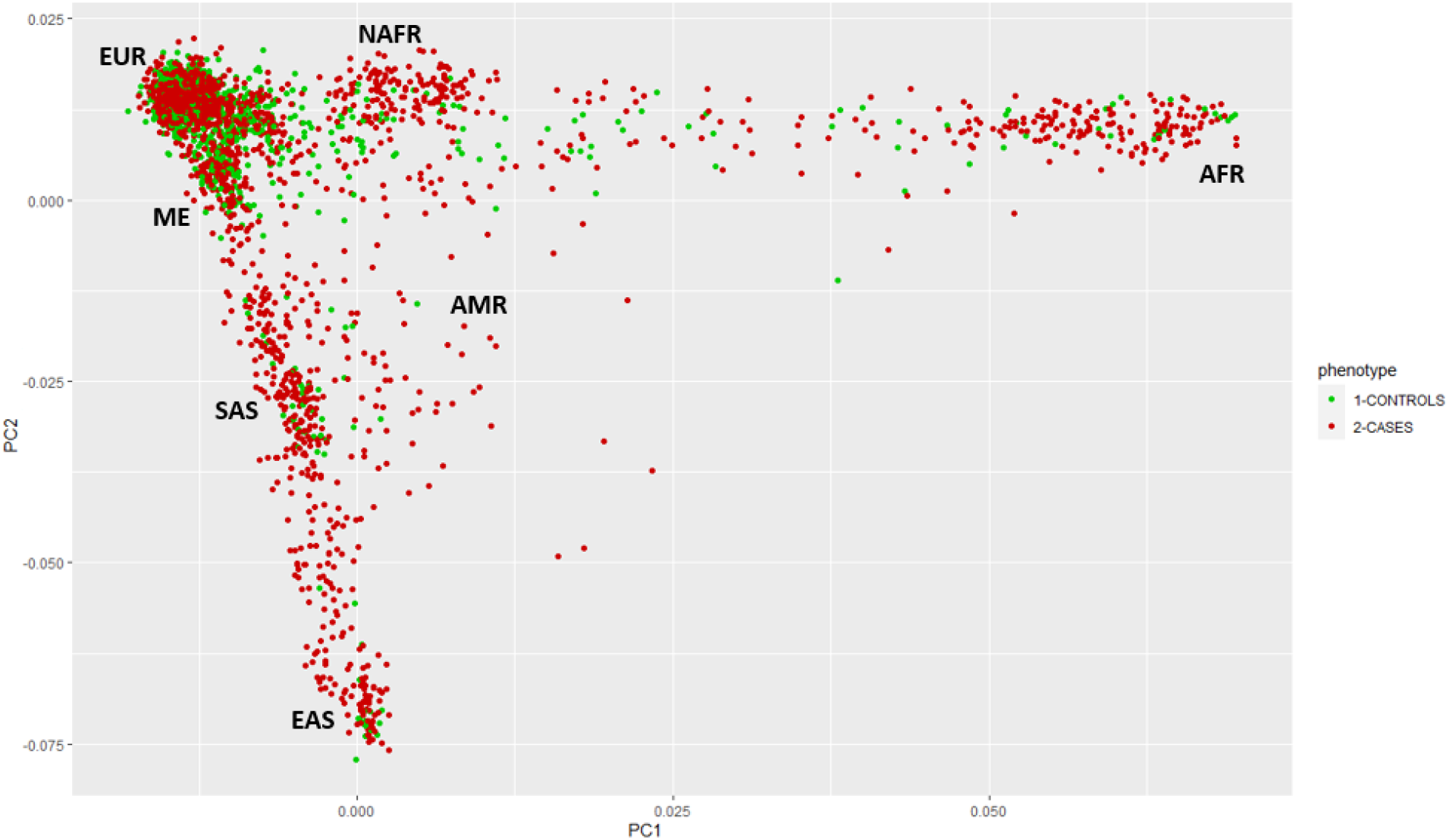
Principal component analysis of patients with life-threatening COVID-19 (red) and controls with asymptomatic or mild infection (green). Principal component analysis (PCA) was performed with PLINK v1.9 software [37] on a pruned subset of ∼14,600 exonic SNPs in linkage equilibrium (maximum r^2^ value for linkage disequilibrium of 0.4 between pairs of SNPs) with a minor allele frequency (MAF) > 1%, call-rate > 99% and *P* value for departure from Hardy-Weinberg equilibrium >10^−5^. Samples were of diverse ethnic origins, including European (EUR), admixed American (AMR), North-African (NAFR), sub-Saharan African (AFR), Middle Eastern (ME), South Asian (SAS) and East Asian (EAS).

### Genome-wide analysis under a co-dominant model

We first performed a GW rare variant burden analysis on the 3,269 patients with life-threatening COVID-19 and 1,373 controls with asymptomatic/mild COVID-19 under a co-dominant model, using nine sets of variants (See Supplemental Methods). The QQ plots for the joint analysis of the samples revealed no systematic deviations from the null hypothesis, and the genomic inflation factors (λ) were close to 1 (Supplemental Table 1). In total, 18,064 genes were analyzed with at least one of the nine variant sets, resulting in an effective number of independent tests (Meff) for the joint analysis of 108,384, giving a Bonferroni-corrected significance threshold of 4.61 × 10^−7^. No gene was found to be of GW significance (see the Manhattan plot in Figure 3A, Supplemental Table 2). The gene with the strongest association was *TREH*, encoding the trehalase enzyme, which hydrolyses trehalose, with rare (GnomAD allele frequency [AF] < 10^−4^) non-synonymous variants associated with a lower risk of life-threatening COVID-19 (OR=0.12[95% CI 0.05-0.28], *P =* 1.9×10^−6^; Supplemental Table 3). In analyses of genes for which rare predicted loss-of-function (pLOF) variants were associated with an increase in the risk of life-threatening COVID-19 (Table 2), the strongest association was that for *NPC2*, for rare (GnomAD AF < 0.01) pLOF variants, with 28 heterozygous carriers among patients (0.9%), and four heterozygous carriers (0.3%) among controls (OR = 5.41 [95% CI 1.8-16.4], *P =* 5.8 × 10^−4^). *NPC2* encodes the Niemann-Pick disease type C2 protein and homozygous LOF mutations of this gene cause Niemann-Pick disease [43]. NPC2 interacts with NPC1, which is also an essential endosomal receptor for the Ebola virus [44,45]. Both NPC1 and NPC2 were implicated in the regulation of SARS-CoV-2 entry in a CRISPR screen[46]. Finally, we analyzed the 19 loci associated with critical pneumonia by GWAS [8,10,12]. None of them showed a significant association (Supplemental Table 4). The GW burden analysis under a dominant model yielded similar conclusions (Supplemental table 3).

**Table 2.** Top results of the genome-wide burden analysis for rare pLOF variants increasing the risk of life-threatening COVID-19 under a co-dominant model.

| Chr | Gene | GnomAD AF threshold | No. carriers of at least one (no. homozygous) pLOF variant |  | Joint analysis |  | Trans-ethnic meta-analysis | Trans-pipeline meta-analysis |
| --- | --- | --- | --- | --- | --- | --- | --- | --- |
|  |  |  | Cases<br>(*n*=3269) | Controls<br>(*n*=1373) | OR[95%CI] | *P* value | *P* value | *P* value |
| 14 | *NPC2* | 0.01 | 28 (0) | 4 (0) | 5.41[1.8-16.4] | 5.8x10<sup>-4</sup> | 2.1x10<sup>-3</sup> | 3.3x10<sup>-4</sup> |
| 3 | *DLEC1* | 0.01 | 56 (0) | 16 (0) | 2.55[1.3-4.9] | 3.6x10<sup>-3</sup> | 0.013 | 4.9x10<sup>-3</sup> |
| 13 | *NEK5* | 0.001 | 16 (0) | 0 (0) | 27.03[0.9-864.2] | 4.0x10<sup>-3</sup> | 1.5x10<sup>-3</sup> | 0.011 |
| 5 | *CCNI2* | 0.01 | 19 (1) | 1 (0) | 7.15[1.2-43.1] | 4.1x10<sup>-3</sup> | 4.1x10<sup>-3</sup> | 5.0x10<sup>-3</sup> |
| 22 | *C22orf29* | 0.001 | 13 (0) | 0 (0) | 15.6[0.8-315.8] | 4.5x10<sup>-3</sup> | 7.9x10<sup>-3</sup> | 4.6x10<sup>-3</sup> |
| 20 | *DLGAP4* | 0.001 | 37 (0) | 3 (0) | 4.35[1.3-14.5] | 4.8x10<sup>-3</sup> | 8.3x10<sup>-3</sup> | 0.011 |

AF: allele frequency
Only genes with a *P* value ≤ 5x10<sup>-3</sup> in the joint analysis and *P* values < 0.05 in trans-ethnic and trans-pipeline meta-analyses are displayed.

**Figure 3.**
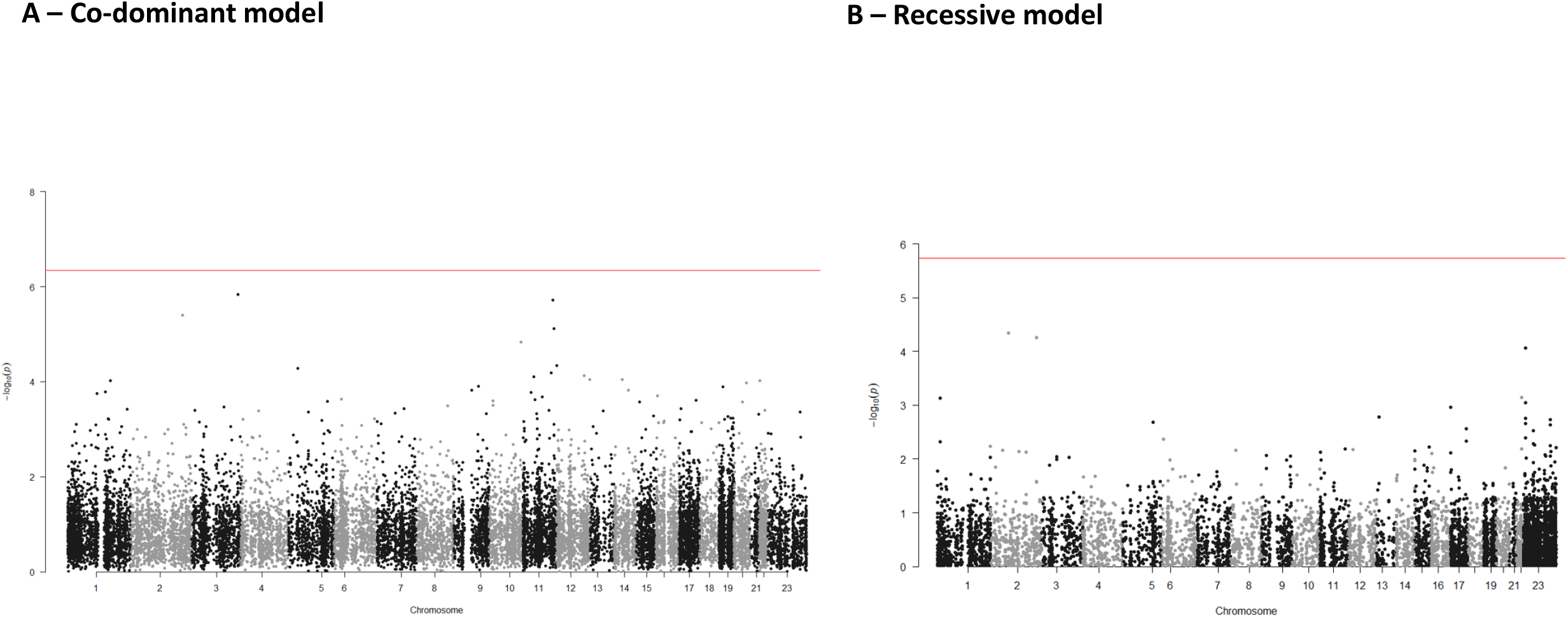
Manhattan plot for the genome-wide burden analysis under the co-dominant (A) and recessive (B) models. The red lines represent the significance threshold after Bonferroni correction accounting for the total number of independent tests (*P =* 4.61 × 10^−7^ under a co-dominant model and 1.85 × 10^−6^ under a recessive model).

### Genome-wide analysis under a recessive model

We then performed a GW screen under a recessive model (autosomal and X-linked). In total, 4,511 genes were analyzed with at least one of the nine variant sets, resulting in 27,066 independent tests, giving a Bonferroni-corrected significance threshold of 1.85 × 10^−6^. No gene reached GW significance (Figure 3B). In analyses of genes with rare variants increasing the risk of life-threatening COVID-19, *TLR7* was, by two orders of magnitude, the most significant gene, with 51 carriers (1.6%) of at least one rare (GnomAD AF < 0.01) missense or pLOF variant in patients versus two carriers (0.1%) in controls (OR = 8.41[95% CI 1.9-35.5], *P =* 8.95×10^−5^) (Table 3). Most of the carriers were male, with only one carrier among the patients and one among the controls being female. The variants carried by the two controls were previously shown to be biochemically neutral [18,28] (Supplemental table 5). The 51 cases carried 33 different variants, 13 of which had been shown to be neutral; 16 were previously shown to be hypomorphic or amorphic [18,28], and four were previously unknown. The four new variants were tested: one was found to be neutral and the other three were deleterious (Supplemental Figure 1). Restricting the analysis to biochemically proven LOF variants (bLOF) decreased the number of carriers (20 cases vs. 0 controls), but the association signal remained highly significant, with a much higher odds ratio (OR = 27.68 [95% CI 1.5-528.7], *P =* 1.08×10^−4^) (Table 3). These findings confirm that *TLR7* is a critical COVID-19 susceptibility locus, responsible for 0.9% of critical cases in male patients.

**Table 3.**
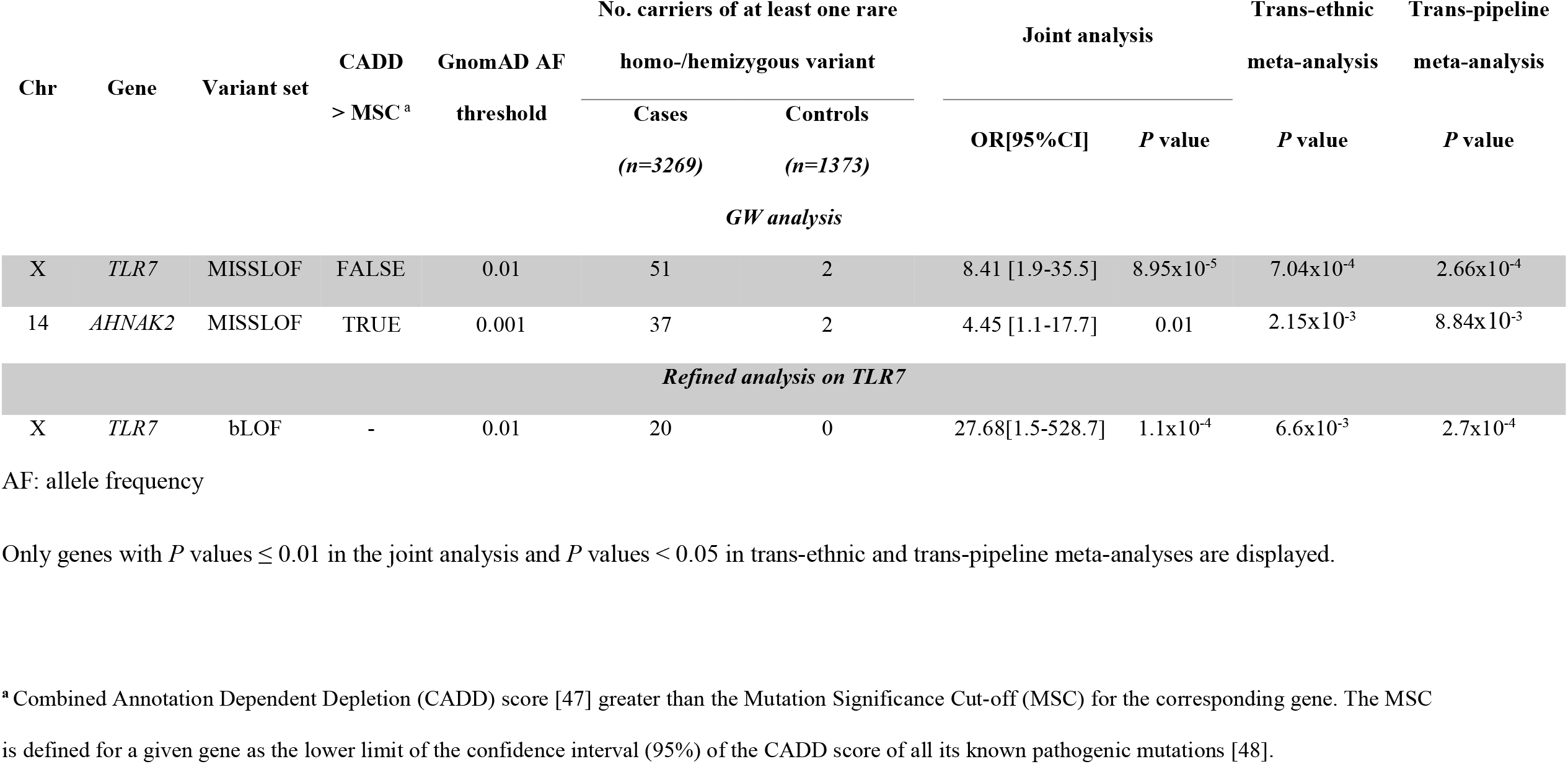
Top results of the genome-wide burden analysis for rare variants increasing the risk of life-threatening COVID-19 under a recessive model.

| Chr | Gene | Variant set | CADD<br>> MSC <sup>a</sup> | GnomAD AF<br>threshold | No. carriers of at least one rare<br>homo-/hemizygous variant |  | Joint analysis |  | Trans-ethnic | Trans-pipeline |
| --- | --- | --- | --- | --- | --- | --- | --- | --- | --- | --- |
|  |  |  |  |  | Cases<br>(n=3269) | Controls<br>(n=1373) | OR[95%CI] | P value | meta-analysis | meta-analysis |
| GW analysis |  |  |  |  |  |  |  |  |  |  |
| X | TLR7 | MISSLOF | FALSE | 0.01 | 51 | 2 | 8.41 [1.9-35.5] | 8.95x10<sup>-5</sup> | 7.04x10<sup>-4</sup> | 2.66x10<sup>-4</sup> |
| 14 | AHNAK2 | MISSLOF | TRUE | 0.001 | 37 | 2 | 4.45 [1.1-17.7] | 0.01 | 2.15x10<sup>-3</sup> | 8.84x10<sup>-3</sup> |
| Refined analysis on TLR7 |  |  |  |  |  |  |  |  |  |  |
| X | TLR7 | bLOF | - | 0.01 | 20 | 0 | 27.68[1.5-528.7] | 1.1x10<sup>-4</sup> | 6.6x10<sup>-3</sup> | 2.7x10<sup>-4</sup> |

AF: allele frequency
Only genes with $P$ values $\leq 0.01$ in the joint analysis and $P$ values $< 0.05$ in trans-ethnic and trans-pipeline meta-analyses are displayed.
<sup>a</sup> Combined Annotation Dependent Depletion (CADD) score [47] greater than the Mutation Significance Cut-off (MSC) for the corresponding gene. The MSC is defined for a given gene as the lower limit of the confidence interval (95%) of the CADD score of all its known pathogenic mutations [48].

### Enrichment in rare pLOF variants at 13 type I IFN-related influenza susceptibility loci

Following on from our initial analysis [14], we also performed a candidate pathway enrichment analysis focusing on the 13 genes involved in Toll-like receptor 3 (TLR3)– and interferon regulatory factor 7 (IRF7)–dependent type I IFN immunity to influenza virus (*IFNAR1, IFNAR2, IRF3, IRF7, IRF9, IKBKG, STAT1, STAT2, TBK1, TICAM1, TLR3, TRAF3* and *UNC93B1*) (Figure 1). We confirmed the significant enrichment in rare (GnomAD AF < 10^−3^) pLOF variants at the 13 loci in patients with critical COVID-19, with 34 carriers among patients versus six among controls (OR = 3.70 [95% CI 1.7-9.5], *P =* 2.1×10^−4^) under a co-dominant model; Table 4). We also estimated this p-value by a simulation study taking at random 13 loci over the whole genome (see supplemental methods). We found an empirical p-value of 2.5×10^−4^. Excluding the 550 cases and 314 controls screened in a previous study [14] resulted in a similar conclusion (OR = 3.21 [95% CI 1.3-8.2], *P* = 5.97×10^−3^). Significant replication was also observed in the trans-ethnic (*P =* 0.01) and the trans-pipeline (*P =* 0.009) analyses. We found that 31 of the 34 carriers of pLOF variants were heterozygous, and three were homozygous: one for a frameshift variant in *IRF7* described in a previous study [14], one for a previously reported deletion spanning 4,394 base pairs in *IFNAR1* [15,18], and one for a previously unknown deletion spanning 6,624 base pairs of *IFNAR1* (Supplemental Table 6). All the homozygous pLOF variants were found in patients. Consequently, the OR for homozygous carriers (OR = 15.79 [95%CI: 1.4-2170.4], *P* = 0.02) was higher than that for heterozygous carriers (OR = 3.11 [95%CI: 1.4-8.6], *P* = 5.2×10^−3^), but both were significant.

**Table 4.** Enrichment in pLOF/bLOF rare variants of genes involved in type I IFN immunity.

| Gene set | Cohort | Model | No. carriers |  | Joint analysis |  | Trans-pipeline<br>meta-analysis | Trans-ethnic<br>meta-analysis |
| --- | --- | --- | --- | --- | --- | --- | --- | --- |
|  |  |  | Cases | Controls | P value | OR[95%CI] | P value | P value |
| 13 genes<sup>a</sup> | Samples independent<br>of [14]<sup>b</sup> | Co-dominant | 25 | 5 | 5.97x10<sup>-3</sup> | 3.21[1.3-8.2] | 9.15x10<sup>-3</sup> | 0.01 |
| 13 genes | Full<sup>c</sup> | Co-dominant | 34 | 6 | 2.13x10<sup>-4</sup> | 3.70[1.7-9.5] | 7.45x10<sup>-4</sup> | 6.52x10<sup>-4</sup> |
| 13 genes | Full | Heterozygous<br>only<sup>d</sup> | 31 | 6 | 5.21x10<sup>-3</sup> | 3.11[1.3-8.6] | 7.88x10<sup>-3</sup> | 5.98x10<sup>-3</sup> |
| 13 genes | Full | Recessive | 3 | 0 | 0.02 | 15.79[1.4-2170.4] | 0.05 | 0.03 |
| 13 genes + *TYK2* | Full | Co-dominant | 37 | 7 | 1.40x10<sup>-4</sup> | 3.30[1.6-7.8] | 5.77x10<sup>-4</sup> | 5.64x10<sup>-4</sup> |
| 13 genes + *TYK2* | Full | Heterozygous<br>only | 32 | 7 | 0.02 | 2.53[1.1-6.6] | 0.03 | 0.02 |
| 13 genes + *TYK2* | Full | Recessive | 5 | 0 | 3.36x10<sup>-3</sup> | 19.65 [2.1-2635.4] | 9.84x10<sup>-3</sup> | 0.03 |
| 13 genes + *TYK2* +<br>bLOF *TLR7* | Full | Co-dominant | 57 | 9 | 1.27x10<sup>-7</sup> | 3.82 [2.0-7.2] | 1.99x10<sup>-7</sup> | 2.20x10<sup>-6</sup> |
| 13genes + *TYK2* +<br>bLOF *TLR7* | Full | Heterozygous<br>only | 32 | 9 | 0.04 | 2.27[1.0-5.2] | 0.04 | 0.02 |
| 13genes + *TYK2* +<br>bLOF *TLR7* | Full | Recessive | 25 | 0 | 4.69x10<sup>-7</sup> | 39.19[5.2-5037.01] | 2.39x10<sup>-6</sup> | 6.66x10<sup>-5</sup> |

<sup>a</sup>*IFNAR1, IFNAR2, IRF3, IRF7, IRF9, IKBKG, STAT1, STAT2, TBK1, TICAM1, TLR3, TRAF3* and *UNC93B1*
<sup>b</sup>2719 patients and 1059 controls newly recruited and not screened in [14]
<sup>c</sup>The full cohort includes 3269 patients and 1373 controls
<sup>d</sup>In this model, only subjects with heterozygous variants are considered as carriers

### Rare pLOF variants of TYK2 and bLOF variants of TLR7

Since the publication of the aforementioned study [14], AR TYK2 deficiency has been reported in children with COVID-19 pneumonia [18]. We identified two patients, already described in another previous study [18], carrying a rare homozygous pLOF variant of *TYK2*, and one patient and one control carrying a rare heterozygous pLOF variant (Supplemental Table 6). Adding these patients to the analysis gave very similar results under a co-dominant model (OR = 3.30[95% CI 1.6-7.8], *P =*1.4×10^−4^) and increased the evidence for association under a recessive model (OR = 19.65[95% CI 2.1-2635.4], *P =*3.4×10^−3^) (Table 4). Analysis of the rare pLOF variants at these 14 loci plus the bLOF variants of *TLR7* revealed highly significant enrichment (OR = 3.82 [95%CI 2.0-7.2], *P* = 1.3×10^−7^ under a co-dominant model). The effect was stronger for homozygous/hemizygous carriers (OR = 39.19 [95%CI 5.2-5037.01], *P* = 4.7×10^−7^) than for heterozygous carriers (OR = 2.27 [95%CI 1.0-5.2], *P* = 0.04), and these two ORs were significantly different (*P* = 0.008). We also found that the 57 patients with critical COVID-19 carrying a rare pLOF or bLOF variant of one of these 15 genes were significantly younger than the remaining 3,212 patients in the cohort (mean age [SD] in years: 43.25 [20.3] vs. 56.0 [17.3] years; *P* = 1.68×10^−5^), consistent with our previous reports that IEIs conferring a predisposition to life-threatening COVID-19 are more frequent in young patients [1,14,28]. Moreover, homozygous/hemizygous carriers were significantly younger than heterozygous carriers (35.2 [20.3] vs. 49.5 [18.2] years, *P* =0.008). Overall, these analyses indicate there is an enrichment in rare pLOF variants at 15 loci involved in type I IFN immunity in patients with critical COVID-19 pneumonia.

### In-frame nonsynonymous variants at the 15 loci

We further screened our cohort for rare in-frame nonsynonymous variants with a GnomAD AF < 10^−3^ at these type I IFN-related susceptibility loci. For the 13 initial loci, the enrichment disappeared when in-frame nonsynonymous variants were added to pLOF variants under a co-dominant model (OR = 1.08 [95%CI 0.9-1.3], *P* = 0.42) (Supplemental Table 7), whereas a non-significant trend persisted under the recessive model (OR = 5.02 [95% CI 0.7-52.7], *P* = 0.06). Focusing exclusively on in-frame variants decreased the strength of this trend considerably, with only eight homozygous carriers among patients and one among controls (OR = 1.14 [0.2-912.5], *P* = 0.68). Adding *TYK2* variants led to similar conclusions (Supplemental table 7). We then added *TLR7* variants and considered the 15 loci together. Under a co-dominant model, the enrichment became non-significant when in-frame nonsynonymous variants were added (OR = 1.15 [1.0-1.4], *P* = 0.09), but it remained significant under a recessive model (OR = 6.54[2.4-24.8], *P* = 5.3×10^−6^; Supplemental Table 7). In analyses considering only rare in-frame homozygous/hemizygous nonsynonymous variants, the effect size was smaller, but the enrichment remained significant (OR = 3.52[1.3-13.3], *P* = 2.8×10^−3^). In total, 41 patients carried a rare homozygous/hemizygous in-frame nonsynonymous variant in one of the 15 loci, and 16 of these variants (carried by 16 patients) were *TLR7* in-frame variants already shown to be bLOF. After excluding the *TLR7* bLOF variants, there was no residual significant enrichment in rare in-frame nonsynonymous variants in patients relative to controls, whatever the genetic model considered.

## Discussion

In this exome-wide gene burden analysis for rare variants underlying critical COVID-19, no gene reached GW statistical significance after accounting for multiple testing. These results are consistent with those of two previous large exome-wide studies including more than 1,000 critical cases and thousands of population-based controls that did not find statistically significant autosomal gene burden associations at stringent significance thresholds accounting for the number of phenotypes and variant sets analyzed [11,21]. However, under a recessive model, the strongest association - although not statistically significant at the GW level - was obtained with the X-linked *TLR7* gene, for which association has consistently been reported across studies [20,28,29], reaching the less conservative exome-wide significance threshold of 2.5×10^−6^ in some of these previous studies [20,21]. It should be stressed that stringent correction for multiple testing, while necessary to avoid false positives, is a conservative strategy, and that the lack of formal statistical significance at a GW level does not preclude biological causality and medical significance. The burden of proof can be provided experimentally via biochemical, virological, and immunological experiments, as we previously did for *TLR7* by showing that biochemically deleterious TLR7 variants blunted the pDC-dependent sensing of SARS-CoV-2 and induction of type I IFN, thereby accounting for ∼1% of critical pneumonia cases in men [28]. Additional genes may be found by restricting the association analysis to variants experimentally proven to be deleterious.

This analysis also confirms our previous findings of an enrichment in rare pLOF variants of 13 genes involved in TLR3- and IRF7-dependent type I IFN immunity to seasonal influenza virus in critical cases relative to controls with mild/asymptomatic infection [14]. These results were strengthened by the addition of *TYK2*, which was recently shown to underlie severe COVID-19 [18,19], and *TLR7*, especially under a recessive model. We found that homozygous/hemizygous carriers of rare pLOF or bLOF variants at the 15 loci had a significantly higher risk of life-threatening COVID-19 than heterozygotes. This is consistent with the generally higher clinical penetrance of recessive than dominant IEI [1]. Overall, 1.7% of the patients with life-threatening COVID-19 carried a rare pLOF or bLOF variant at one of the 15 loci, these variants being homozygous/hemizygous in 0.8%. The study of in-frame nonsynonymous variants might increase this proportion, but would require the experimental characterization of all these variants. Indeed, in analyses restricted to rare in-frame nonsynonymous variants, we detected no significant enrichment in patients relative to controls. This result is not surprising, as we showed in a previous study [14] that less than 15% of the rare in-frame nonsynonymous variants at the 13 initially studied loci carried by cases were bLOF variants, whereas all the pLOF variants were found to be bLOF. Similar results were obtained for *TLR7*, with only 10 of 108 (9.2%) in-frame nonsynonymous variants observed in GnomAD being bLOF [28]. This high proportion of neutral variants strongly affects the power of burden tests and highlights the need for the experimental characterization of variants.

We also showed that patients carrying rare pLOF or bLOF variants at these 15 type I IFN-related genes were significantly younger than the remaining patients (mean age [SD] in years: 43.3 [20.3] vs. 56.0 [17.3] years). This was particularly true for carriers of a homozygous/hemizygous rare pLOF or bLOF variant (35.2 [20.3] years), potentially accounting for the lack of replication of this finding by other studies including older patients [11,20–22]. Consistent with this result, we recently found that ∼10% of children hospitalized for COVID-19 pneumonia carry recessive inborn errors of type I IFN immunity [18]. In addition, older patients are more likely to carry auto-Abs against type I IFN, and contrary to other previous studies, we excluded patients carrying such antibodies from the analysis. None of the 234 patients with critical COVID-19 excluded from this study due to the presence of auto-Abs against type I IFN carried a rare pLOF variant of the 15 genes. Hence, samples in which the vast majority of patients are over the age of 60 years and of unknown status for auto-Abs against type I IFNs would have much reduced power to identify these rare inborn errors of type I IFN immunity. In conclusion, rare autosomal inborn errors of type I IFN-dependent immunity to influenza viruses can underlie critical forms of COVID-19, especially in subjects below 60 years of age, in addition to X-linked TLR7 deficiency. The search for additional rare mutations conferring a strong predisposition to life-threatening COVID-19 should focus on young patients with critical COVID-19 without auto-Abs against type I IFNs.

## Data Availability

Data supporting the findings of this study are available within the manuscript and supplemental
files.

## Declarations

### Ethics approval and consent to participate

All the enrolled participants provided written informed consent for participation and were recruited through protocols conforming to local ethics requirements. For patients enrolled in the French COVID cohort (ClinicalTrials.gov NCT04262921), ethics approval was obtained from the Comité de Protection des Personnes Ile De France VI (ID RCB, 2020-A00256-33) or the Ethics Committee of Erasme Hospital (P2020/203). For participants enrolled in the COV-Contact study (ClinicalTrials.gov NCT04259892), ethics approval was obtained from the CPP IDF VI (ID RCB, 2020-A00280-39). For patients enrolled in the Italian cohort, ethics approval was obtained from the University of Milano-Bicocca School of Medicine, San Gerardo Hospital, Monza–Ethics Committee of the National Institute of Infectious Diseases Lazzaro Spallanzani (84/2020) (Italy), and the Comitato Etico Provinciale (NP 4000–Studio CORONAlab). STORM-Health care workers were enrolled in the STudio OsseRvazionale sullo screening dei lavoratori ospedalieri per COVID-19 (STORM-HCW) study, with approval from the local institutional review board (IRB) obtained on June 18, 2020. Patients and relatives from San Raffaele Hospital (Milan) were enrolled in COVID-BioB/Gene-COVID protocols and, for additional studies, TIGET-06, with the approval of the local ethics committee. Patients and relatives from Rome were enrolled in Protocol no. 50/20 (Tor Vergata University Hospital). Informed consent was obtained from each patient. For the patients enrolled in the COVIDeF Study Group (ClinicalTrials.gov NCT04352348), ethics approval was obtained from the Comité de Protection des Personnes Ile de France XI (ID RCB, 2020-A00754-35). For patients enrolled in Spain, the study was approved by the Committee for Ethical Research of the Infanta Leonor University Hospital, code 008-20; the Committee for Ethical Research of the 12 de Octubre University Hospital, code 16/368; the Bellvitge University Hospital, code PR127/20; the University Hospital of Gran Canaria Dr. Negrín, code 2020-200-1 COVID-19; and the Vall d’Hebron University Hospital, code PR(AMI)388/2016. Anonymized samples were sequenced at the National Institute of Allergy and Infectious Diseases (NIAID) through the Uniformed Services University of the Health Sciences (USUHS)/the American Genome Center (TAGC) under nonhuman subject research conditions; no additional IRB consent was required at the National Institutes of Health (NIH). For patients enrolled in the Swedish COVID cohort, ethics approval was obtained from the Swedish Ethical Review Agency (2020-01911 05).

## Data and materials availability

Data supporting the findings of this study are available within the manuscript and supplemental files. Patients were not consented to share the raw WES/WGS data files beyond the research and clinical teams. NIH patient data are available under dbGaP submission phs002245.v1.

## Author contributions

D Matuozzo, ET, AM, JM, YS, YZ, A Bolze, MC, BM, P Zhang, LA and AC performed computational analysis. D Matuozzo, AG, P Bastard, TA, LB, I Meyts, SYZ, A Puel, SBD, BB, EJ and QZ performed or supervised experiments, generated and analyzed data, and contributed to the manuscript by providing figures and tables. P Bastard, FB, HA, AAT, AA, IAD, LMA, RAA, AAA, GA, P Bergman, SB, YTB, IGB, OCM, SC, PC, GC, KC, RC, CAN, LEZ, CF, PKG, MG, FH, RH, SH, LH, NH, AK, SK, CK, RLL, JLF, D Mansouri, JMP, OMA, I Migeotte, PEM, GM, AMN, GN, AN, TO, FP, QPH, RP, LPS, DEP, CP, A Pujol, LFR, JGR, CRG, JR, PRQ, MS, A Sobh, PSP, YTL, IT, CT, JT, MZ, P Zawadzki, SZAM, HBF, MJB, SNC, MAC, CLD, JF, JRH, YLL, RPL, TM, THM, HVB, AL, MV, A Boland, JFD, FM, ST, GG, FT, PH, LDN and HCS evaluated and recruited patients and /or controls. CRG, A Schlüter, MS, MZ, P Zawadzki, SZAM, HBF, MJB, SNC, MAC, CLD, JF, JRH, YLL, RPL, TM, THM, HVB, AL, MV, A Boland, JFD, RN and AKK performed sequencing. D Matuozzo, BB, JLC, QZ, LA and AC wrote the manuscript. JLC, QZ, LA and AC supervised the project. All the authors edited the manuscript and approved its final version.

## Acknowledgements and fundings

We thank the patients and their families for agreeing to participate in our research. The Laboratory of Human Genetics of Infectious Diseases is supported by the Howard Hughes Medical Institute, the Rockefeller University, the St. Giles Foundation, the National Institutes of Health (NIH) (R01AI088364 and R01AI63029), the National Center for Advancing Translational Sciences (NCATS), NIH Clinical and Translational Science Award (CTSA) program (UL1 TR001866), a Fast Grant from Emergent Ventures, Mercatus Center at George Mason University, the Yale Center for Mendelian Genomics and the GSP Coordinating Center funded by the National Human Genome Research Institute (NHGRI) (UM1HG006504 and U24HG008956), the Yale High Performance Computing Center (S10OD018521), the Fisher Center for Alzheimer’s Research Foundation, the JPB Foundation, the Meyer Foundation, the French National Research Agency (ANR) under the “Investments for the Future” program (ANR-10-IAHU-01), the Integrative Biology of Emerging Infectious Diseases Laboratory of Excellence (ANR-10-LABX-62-IBEID), the French Foundation for Medical Research (FRM) (EQU201903007798), the ANR GenMISC (ANR-21-COVR-039), the ANRS-COV05, ANR GENVIR (ANR-20-CE93-003) ANR AABIFNCOV (ANR-20-CO11-0001) projects, the ANR-RHU program (ANR-21-RHUS-08), the European Union’s Horizon 2020 research and innovation program under grant agreement No. 824110 (EASI-genomics), the HORIZON-HLTH-2021-DISEASE-04 program under grant agreement 01057100 (UNDINE), the Square Foundation, Grandir - Fonds de solidarité pour l’enfance, Fondation du Souffle, the SCOR Corporate Foundation for Science, The French Ministry of Higher Education, Research, and Innovation (MESRI-COVID-19), Institut National de la Santé et de la Recherche Médicale (INSERM), REACTing-INSERM and the University of Paris Cité. The study was supported by the ORCHESTRA project, which has received funding from the European Union’s Horizon 2020 research and innovation program under grant agreement No 10101616. P Bastard was supported by the MD-PhD program of the Imagine Institute (with the support of the Fondation Bettencourt-Schueller). The French COVID Cohort study group was sponsored by INSERM and supported by the REACTing consortium and by a grant from the French Ministry of Health (Grant PHRC 20-0424). The Cov-Contact Cohort was supported by the REACTing consortium, the French Ministry of Health, and the European Commission (Grant RECOVER WP 6). The COVIDeF study group was supported by the French Ministry of Health, Fondation AP-HP et Programme Hospitalier de Recherche Clinique (PHRC COVID-19-20-0048). H.C.S is supported by the Intramural Research Program of the National Institute of Allergy and Infectious Diseases, NIH. G.N. and A.N. are supported by Regione Lazio (Research Group Projects 2020) No. A0375-2020-36663, GecoBiomark. I Meyts is a Senior Clinical Investigator at the Research Foundation – Flanders, and is supported by the CSL Behring Chair of Primary Immunodeficiencies, by the KU Leuven C1 Grant C16/18/007, by a VIB GC PID Grant, by the FWO Grants G0C8517N, G0B5120N and G0E8420N and by the Jeffrey Modell Foundation. This project has received funding from the European Research Council (ERC) under the European Union’s Horizon 2020 research and innovation program (grant agreement no. 948959). This work is supported by the Swiss National Science Foundation (grant # 310030L_197721 to JF). This work is supported by ERN-RITA. The Canarian Sequencing Hub is funded by Instituto de Salud Carlos III (COV20_01333, and COV20_01334, and PI20/00876) and Spanish Ministry of Science and Innovation (RTC-2017-6471-1; AEI/FEDER, UE), co-financed by the European Regional Development Funds, “A way of making Europe” from the European Union, and Cabildo Insular de Tenerife (CGIEU0000219140 and “Apuestas científicas del ITER para colaborar en la lucha contra la COVID-19”). This work was funded, at least in part, by grant AJF202059 from Al Jalila Foundation, Dubai, United Arab Emirates. Sample processing at IrsiCaixa was possible thanks to the crowdfunding initiative YoMeCorono. We thank I Erkizia, E Grau, M Massanella, and J Guitart from the IrsiCaixa and Hospital Germans Trias i Pujol (Badalona, Spain) for sample collection, handling and processing. See Supplemental Acknowledgments for the list of consortia members.

