## Supplemental material for "Rare predicted loss-of-function variants of type I IFN immunity genes are associated with life-threatening COVID-19"

**Supplemental Methods:**

***Sequencing***

The whole-exome (N= 2003 cases and 866 controls) or whole-genome (N=1266 cases and 507 controls) was sequenced at several sequencing centers, including the Genomics Core Facility of the Imagine Institute (Paris, France), the Yale Center for Genome Analysis (USA), Macrogen (USA), Psomagen (USA), the New-York Genome Center (NY, USA), TAGC (USUHS, Bethesda, USA), MNM Bioscience (Poland), Invitae (San Francisco, USA), the Genomic Sequencing Platform Seqoia (France), the Centre National de Recherche en Génomique Humaine (CNRGH, Evry, France), the Genomics Division-ITER of the Canarian Health System sequencing hub (Canary Islands, Spain), the AlJalila Genomics Center (Dubai). Libraries for WES were generated with the Twist and Twist Plus Human Core Exome Kit, the xGen Exome Research Panel from Integrated DNA Technologies (IDT; xGen V1 and V2), Agilent SureSelect (Clinical Research exome V2, Human All Exon V6 and V7) panels, the SeqCap EZ MedExome Kit from Roche, the Nextera Flex for Enrichment-Exome kit, the Illumina TruSeq Exome panel and WES custom target enrichlent probes. Massively parallel sequencing was performed on HiSeq 4000, HiSeq 2500, NextSeq 550 or NovaSeq 6000 systems (Illumina). For 3363 samples (2493 critical cases and 870 controls,), the FASTQ files were centralized and processed in the laboratory of Human Genetics of Infectious Diseases (HGID). Raw reads were aligned and mapped to the human reference genome assembly hg19 – NCBI build 37 using the Burrows-Wheeler Aligner (BWA). Post alignment processing procedures from Genome Analysis Software Kit (GATK version 3.4-46) best-practice pipeline were applied to minimize eventual artifacts that may affect the quality of WES/WGS data. PCR duplicates were removed with Picard tools (broadinstitute.github.io/picard/). The GATK base quality score recalibrator (BQSR) was applied to correct sequencing artifacts. Individual genomic variant call files (gVCF) were generated with GATK HaplotypeCaller, and joint genotyping was performed with GATK GenotypeGVCFs in the interval intersecting all the main WES capture kits of ±50 bp. We also searched for rare homozygous or hemizygous deletions from the NGS data using HMZDelFinder-opt [1]. For the remaining 1280 samples (777 cases and 503 controls,), raw data were processed separately by each sequencing hub using either GATK best-practice pipeline or Illumina DRAGEN pipeline. For those samples, only single sample VCF files were centralized in the laboratory of Human Genetics of Infectious Diseases. VCF files were merged together and with the multi-sample VCF generated by the HGID pipeline using BCFtools v1.9 (http://github.com/samtools/bcftools), assuming that genotypes at missing sites were homozygous reference. Only variants located in the interval intersecting all the main WES capture kits ±50 bp were kept.

### *Quality Control*

Per-sample quality control (QC) metrics (mean variant depth, number of indels, transitions/transversions rate, number of heterozygous, reference/non reference homozygous ratio) were generated using BCFtools v1.9 (http://github.com/samtools/bcftools) and samples outlier on the QC metric distributions were excluded. We estimated the kinship coefficient between all pairs of samples using the related option of the king software and excluded first degree relatives. Sex was inferred from the coverage of the X and Y chromosomes and samples with inconsistencies between the reported and inferred sex were excluded. Sample genotypes with a coverage of <8×, a genotype quality of <20, or a ratio of reads for the less covered allele (reference or variant allele) over the total number of reads covering the position (minor read ratio) of <20% for heterozygous calls were filtered out. We excluded variant sites (i) with a call rate of <50% in gnomAD genomes and exomes, (ii) with a non-PASS filter in the gnomAD database, (iii) falling in low-complexity or decoy regions, (iv) that were multiallelic with more than four alleles,(v) spanning more than 20 nucleotides, (vi) with more than 5% missing genotypes in our cohort with raw data available, (vii) with differential missingness between cases and controls (P-value $<10^{-4}$), (viii) blacklisted using our blacklist procedure [2]. We generated four blacklists (i.e. variants absent from Gnomad v2.1 while present at frequency > 1% in our dataset) according to the type of data available (FASTQ vs VCF only) and the type of sequencing (WES vs WGS). Variants falling in at least one of the four blacklists were excluded. For variants identified in the 13 type I IFN-related influenza susceptibility loci, manual review of aligned reads for variant confirmation was done with The Integrative Genomics Viewer [3].

### *Variant annotation*

Variant effects were predicted with the Ensembl Variant Effect Predictor and the Ensembl GRCh37.75 reference database, retaining the most deleterious annotation per gene obtained from Ensembl protein-coding transcripts overlapping with RefSeq transcripts. The Combined Annotation Dependent Depletion (CADD) score[4] was used to predict the potential deleteriousness of the variants. However, the CADD score for known pathogenic mutations is highly variable from one gene to another and it may be useful to consider specific gene thresholds rather than a fixed threshold; to do so, we used the MSC ("Mutation Significance Cut-off") which is defined for a given gene as the lower limit of the confidence interval (95%) of the CADD score of all its known pathogenic mutations [5]. Thus, variants with a CADD score higher than the MSC are more likely to lead to an altered phenotype than variants with a CADD lower than the MSC. In addition, allele frequencies in exomes and genomes were retrieved for each variant from the Genome Aggregation Database (GnomAD v2.1).

### *Simulation study*

We performed a simulation study to assess the empirical p-value for the test of enrichment in rare pLOF variants in cases at 13 selected loci. In each of the 100,000 replicates, we selected at random 13 loci among the 18018 loci available at the genome level, and tested for an enrichment of pLOF variants with frequency < $10^{-3}$ at those 13 randomly selected loci in our 3269 critical cases as compared with our 1373 infected controls. Genes for which more than 1% of individuals carried a pLOF variant were excluded from the list of available loci. The analyses were performed by means of LRT with Firth's bias correction under a codominant model of inheritance, and were adjusted on sex, age (in years) and five first PCs of the PCA. To estimate the empirical p-value, we extracted the number of replicates with enrichment of pLOF variants in cases that gave a lower p-value than the one observed for the enrichment in rare pLOF variants at 13 type I IFN-related influenza susceptibility loci ($P = 2.1x10^{-4}$), and divided it to the total number of replications (100,000).

### *Luciferase reporter assay*

We tested the *TLR7* variants as previously described [6]. Briefly, HEK293T cells, which have no endogenous TLR7 expression, were transfected with the pCMV6 vector bearing WT or variant TLR7 (50 ng), the reporter construct pGL4.32 (100 ng), and an expression vector for Renilla luciferase (10 ng), with the X-tremeGENE 9 DNA Transfection Reagent kit (Sigma-Aldrich). The pGL4.32 (luc2P/NF-κB–RE/Hygo) (Promega) reporter vector contains five copies of the NF-κB–responsive element (NF-κB–RE) linked to the luciferase reporter gene luc2P. After 24 hours, the transfected cells were left unstimulated or were stimulated with R848

(1 μg/ml; resquimod), for activation via TLR7/8 (Invivogen), or R837 (5 μg/ml; imiquimod) (Invivogen), or CL264 (5 μg/ml; Invivogen), human TLR7-specific agonists, for 24 hours. Relative luciferase activity was then determined by normalizing the values against the firefly:Renilla luciferase signal ratio.

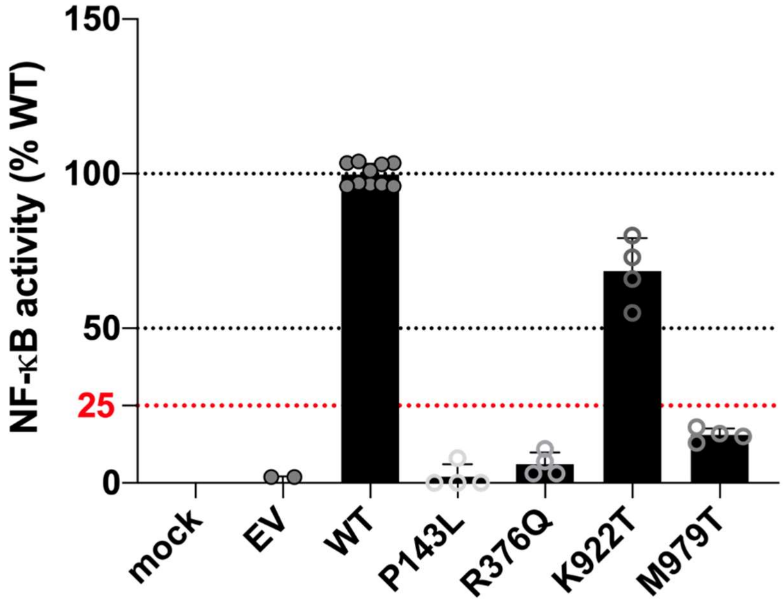


**Supplemental Figure S1.** Luciferase assay on HEK293T cells transfected with the pGL4.32 luciferase reporter construct and an expression vector for *Renilla* luciferase together with no vector (mock), EV, WT, or 4 *TLR7* variants found in our cohort. After 24 hours, transfected cells were left untreated or were treated by incubation with 1 µg/mL R848 for 24 hours. These data were established from two independent experiments. The *y*-axis represents NF-κB transcriptional activity as a percentage of the WT. The *x*-axis indicates the alleles used fortransfection.

**List of consortia members**

**Members of COVID Human Genetic Effort:** Laurent Abel[1], Alessandro Aiuti[2], Saleh Al-Muhsen[3], Fahd Al-Mulla[4], Mark S. Anderson[5], Evangelos Andreakos[6], Andrés A. Arias[7], Hagit Baris Feldman[8], Alexandre Belot[9], Catherine M. Biggs[10], Dusan Bogunovic[11], Alexandre Bolze[12], Anastasiia Bondarenko[13], Ahmed A. Bousfiha[14], Petter Brodin[15], Yenan Bryceson[16], Carlos D. Bustamante[17], Manish J. Butte[18], Giorgio Casari[19], Samya Chakravorty[20], John Christodoulou[21], Antonio Condino-Neto[22], Stefan N. Constantinescu[23], Megan A. Cooper[24], Clifton L. Dalgard[25], Murkesh Desai[26], Beth A. Drolet[27], Jamila El Baghdadi[28], Sara Espinosa-Padilla[29], Jacques Fellay[30], Carlos Flores[31], José Luis Franco[7], Antoine Froidure[32], Peter K. Gregersen[33], Filomeen Haerynck[34], David Hagin[35], Rabih Halwani[36], Lennart Hammarström[37], James R. Heath[38], Sarah E. Henrickson[39], Elena W. Y. Hsieh[40], Eystein Husebye[41], Kohsuke Imai[42], Yuval Itan[43], Erich D. Jarvis[44], Timokratis Karamitros[45], Kai Kisand[46], Cheng-Lung Ku[47], Yu-Lung Lau[48], Yun Ling[49], Carrie L. Lucas[50], Tom Maniatis[51], Davood Mansouri[52], László Maródi[53], Isabelle Meyts[54], Joshua D. Milner[55], Kristina Mironska[56], Trine H. Mogensen[57], Tomohiro Morio[58], Lisa F. P. Ng[59], Luigi D. Notarangelo[60], Antonio Novelli[61], Giuseppe Novelli[62], Cliona O'Farrelly[63], Satoshi Okada[64], Tayfun Ozcelik[65], Qiang Pan-Hammarström[37], Rebeca Perez de Diego[66], Anna M. Planas[67], Carolina Prando[68], Aurora Pujol[69], Lluis Quintana-Murci[70], Laurent Renia[59], Igor Resnick[71], Carlos Rodríguez-Gallego[72], Vanessa Sancho-Shimizu[73], Anna Sediva[74], Mikko R. J. Seppänen[75], Mohammed Shahrooei[76], Anna Shcherbina[77], Ondrej Slaby[78], Andrew L. Snow[79], Pere Soler-Palacín[80], András N. Spaan[81], Ivan Tancevski[82], Stuart G. Tangye[83], Ahmad Abou Tayoun[84], Sathishkumar Ramaswamy[84], Stuart E. Turvey[85], Furkan Uddin[86], Mohammed J. Uddin[87], Diederik van de Beek[88], Donald C. Vinh[89], Horst von Bernuth[90], Mayana Zatz[91], Pawel Zawadzki[92], Helen C. Su[60], Jean-Laurent Casanova[93]

[1]INSERM U1163, University of Paris, Imagine Institute, Paris, France. [2]San Raffaele Telethon Institute for Gene Therapy, IRCCS Ospedale San Raffaele, and Vita Salute San Raffaele University, Milan, Italy. [3]Immunology Research Laboratory, Department of Pediatrics, College of Medicine and King Saud University Medical City, King Saud University, Riyadh, Saudi Arabia. [4]Dasman Diabetes Institute, Department of Genetics and Bioinformatics, Dasman, Kuwait. [5]Diabetes Center, University of California, San Francisco, San Francisco, CA, USA. [6]Laboratory of Immunobiology, Center for Clinical, Experimental Surgery and Translational Research, Biomedical Research Foundation of the Academy of Athens, Athens, Greece. [7]Group of Primary Immunodeficiencies, Universidad de Antioquia UdeA, Medellín, Colombia. [8]Genetics Institute, Tel Aviv Sourasky Medical Center and Sackler Faculty of Medicine, Tel Aviv University, Tel Aviv, Israel. [9]Pediatric Nephrology, Rheumatology, Dermatology, HFME, Hospices Civils de Lyon, National Referee Centre RAISE, and INSERM U1111, Université de Lyon, Lyon, France. [10]Department of Pediatrics, British Columbia Children's Hospital, University of British Columbia, Vancouver, BC, Canada. [11]Icahn School of Medicine at Mount Sinai, New York, NY, USA. [12]Helix, San Mateo, CA, USA. [13]Shupyk National Medical Academy for Postgraduate Education, Kiev, Ukraine. [14]Clinical Immunology Unit, Department of Pediatric Infectious Disease, CHU Ibn Rushd and LICIA, Laboratoire d'Immunologie Clinique, Inflammation et Allergie, Faculty of Medicine and Pharmacy, Hassan II University, Casablanca, Morocco. [15]SciLifeLab, Department Of Women's and Children's Health, Karolinska Institutet, Stockholm, Sweden. [16]Department of Medicine, Center for Hematology and Regenerative Medicine, Karolinska Institutet, Stockholm, Sweden. [17]Stanford University, Stanford, CA, USA. [18]Division of Immunology, Allergy, and Rheumatology, Department of Pediatrics and the Department of Microbiology, Immunology, and Molecular Genetics, University of California, Los Angeles, Los Angeles, CA, USA. [19]Clinical Genomics, IRCCS San Raffaele Scientific Institute and Vita-Salute San Raffaele University, Milan,

Italy. [20]Department of Pediatrics and Children's Healthcare of Atlanta, Emory University, Atlanta, GA, USA. [21]Murdoch Children's Research Institute and Department of Paediatrics, University of Melbourne, Melbourne, VIC, Australia. [22]Department of Immunology, Institute of Biomedical Sciences, University of São Paulo, São Paulo, Brazil. [23]de Duve Institute and Ludwig Cancer Research, Brussels, Belgium. [24]Washington University School of Medicine, St. Louis, MO, USA. [25]Department of Anatomy, Physiology and Genetics, Uniformed Services University of the Health Sciences, Bethesda, MD, USA. [26]Bai Jerbai Wadia Hospital for Children, Mumbai, India. [27]School of Medicine and Public Health, University of Wisconsin, Madison, WI, USA. [28]Genetics Unit, Military Hospital Mohamed V, Rabat, Morocco. [29]Instituto Nacional de Pediatria (National Institute of Pediatrics), Mexico City, Mexico. [30]School of Life Sciences, Ecole Polytechnique Fédérale de Lausanne, Lausanne, Switzerland; Precision Medicine Unit, Lausanne University Hospital and University of Lausanne, Lausanne, Switzerland. [31]Genomics Division, Instituto Tecnológico y de Energías Renovables (ITER), Santa Cruz de Tenerife, Spain; Research Unit, Hospital Universitario N.S. de Candelaria, Santa Cruz de Tenerife, Spain; Instituto de Tecnologías Biomédicas (ITB), Universidad de La Laguna, San Cristóbal de La Laguna, Spain; CIBER de Enfermedades Respiratorias, Instituto de Salud Carlos III, Madrid, Spain. [32]Pulmonology Department, Cliniques Universitaires Saint-Luc; Institut de Recherche Expérimentale et Clinique (IREC), Université Catholique de Louvain, Brussels, Belgium. [33]Feinstein Institute for Medical Research, Northwell Health USA, Manhasset, NY, USA. [34]Department of Paediatric Immunology and Pulmonology, Centre for Primary Immunodeficiency Ghent (CPIG), PID Research Laboratory, Jeffrey Modell Diagnosis and Research Centre, Ghent University Hospital, Ghent, Belgium. [35]Genetics Institute Tel Aviv Sourasky Medical Center, Tel Aviv, Israel. [36]Sharjah Institute of Medical Research, College of Medicine, University of Sharjah, Sharjah, United Arab Emirates. [37]Department of Biosciences and Nutrition, Karolinska Institutet, Stockholm, Sweden. [38]Institute for Systems Biology, Seattle, WA, USA. [39]Department of Pediatrics, Division of Allergy Immunology, Children's Hospital of Philadelphia, Philadelphia, PA, USA; Department of Microbiology, Perelman School of Medicine, University of Pennsylvania, Philadelphia, PA, USA. [40]Departments of Pediatrics, Immunology and Microbiology, University of Colorado, School of Medicine, Aurora, CO, USA. [41]Department of Medicine, Haukeland University Hospital, Bergen, Norway. [42]Department of Community Pediatrics, Perinatal and Maternal Medicine, Tokyo Medical and Dental University (TMDU), Tokyo, Japan. [43]Institute for Personalized Medicine, Icahn School of Medicine at Mount Sinai, New York, NY, USA; Department of Genetics and Genomic Sciences, Icahn School of Medicine at Mount Sinai, New York, NY, USA. [44]Laboratory of Neurogenetics of Language and Howard Hughes Medical Institute, Rockefeller University, New York, NY, USA. [45]Bioinformatics and Applied Genomics Unit, Hellenic Pasteur Institute, Athens, Greece. [46]Molecular Pathology, Department of Biomedicine, Institute of Biomedicine and Translational Medicine, University of Tartu, Tartu, Estonia. [47]Chang Gung University, Taoyuan County, Taiwan. [48]Department of Paediatrics and Adolescent Medicine, University of Hong Kong, Hong Kong, China. [49]Shanghai Public Health Clinical Center, Fudan University, Shanghai, China. [50]Department of Immunobiology, Yale University School of Medicine, New Haven, CT, USA. [51]Columbia University Zuckerman Institute, New York, NY, USA. [52]Department of Clinical Immunology and Infectious Diseases, National Research Institute of Tuberculosis and Lung Diseases, Clinical Tuberculosis and Epidemiology Research Center, National Research Institute of Tuberculosis and Lung Diseases (NRITLD), Masih Daneshvari Hospital, Shahid Beheshti University of Medical Sciences, Tehran, Iran. [53]Primary Immunodeficiency Clinical Unit and Laboratory, Department of Dermatology, Venereology and Dermatooncology, Semmelweis University, Budapest, Hungary. [54]Department of Pediatrics, University Hospitals Leuven; KU Leuven, Department of

Microbiology, Immunology and Transplantation; Laboratory for Inborn Errors of Immunity, KU Leuven, Leuven, Belgium. [55]Department of Pediatrics, Columbia University Irving Medical Center, New York, NY, USA. [56]University Clinic for Children's Diseases, Department of Pediatric Immunology, Medical Faculty, University "St.Cyril and Methodij," Skopje, North Macedonia. [57]Department of Biomedicine, Aarhus University, Aarhus, Denmark. [58]Tokyo Medical and Dental University Hospital, Tokyo, Japan. [59]A*STAR Infectious Disease Labs, Agency for Science, Technology and Research, Singapore, Singapore; Lee Kong Chian School of Medicine, Nanyang Technology University, Singapore, Singapore. [60]National Institute of Allergy and Infectious Diseases, National Institutes of Health, Bethesda, MD, USA. [61]Laboratory of Medical Genetics, IRCCS Bambino Gesù Children's Hospital, Rome, Italy. [62]Department of Biomedicine and Prevention, Tor Vergata University of Rome, Rome, Italy. [63]Comparative Immunology Group, School of Biochemistry and Immunology, Trinity Biomedical Sciences Institute, Trinity College Dublin, Dublin, Ireland. [64]Department of Pediatrics, Graduate School of Biomedical and Health Sciences, Hiroshima University, Hiroshima, Japan. [65]Department of Molecular Biology and Genetics, Bilkent University, Bilkent-Ankara, Turkey. [66]Laboratory of Immunogenetics of Human Diseases, Innate Immunity Group, IdiPAZ Institute for Health Research, La Paz Hospital, Madrid, Spain. [67]IIBB-CSIC, IDIBAPS, Barcelona, Spain. [68]Faculdades Pequeno Príncipe, Instituto de Pesquisa Pelé Pequeno Príncipe, Curitiba, Brazil. [69]Neurometabolic Diseases Laboratory, Bellvitge Biomedical Research Institute (IDIBELL), L'Hospitalet de Llobregat, Barcelona, Spain; Catalan Institution of Research and Advanced Studies (ICREA), Barcelona, Spain; Center for Biomedical Research on Rare Diseases (CIBERER), ISCIII, Barcelona, Spain. [70]Human Evolutionary Genetics Unit, CNRS U2000, Institut Pasteur, Paris, France; Human Genomics and Evolution, Collège de France, Paris, France. [71]University Hospital St. Marina, Varna, Bulgaria. [72]Department of Immunology, University Hospital of Gran Canaria Dr. Negrín, Canarian Health System, Las Palmas de Gran Canaria, Spain; Department of Clinical Sciences, University Fernando Pessoa Canarias, Las Palmas de Gran Canaria, Spain. [73]Department of Paediatric Infectious Diseases and Virology, Imperial College London, London, UK; Centre for Paediatrics and Child Health, Faculty of Medicine, Imperial College London, London, UK. [74]Department of Immunology, Second Faculty of Medicine Charles University, V Uvalu, University Hospital in Motol, Prague, Czech Republic. [75]Adult Immunodeficiency Unit, Infectious Diseases, Inflammation Center, University of Helsinki and Helsinki University Hospital, Helsinki, Finland; Rare Diseases Center and Pediatric Research Center, Children's Hospital, University of Helsinki and Helsinki University Hospital, Helsinki, Finland. [76]Saeed Pathobiology and Genetics Lab, Tehran, Iran; Department of Microbiology and Immunology, Clinical and Diagnostic Immunology, KU Leuven, Leuven, Belgium. [77]Department of Immunology, Dmitry Rogachev National Medical Research Center of Pediatric Hematology, Oncology and Immunology, Moscow, Russia. [78]Central European Institute of Technology and Department of Biology, Faculty of Medicine, Masaryk University, Brno, Czech Republic. [79]Department of Pharmacology and Molecular Therapeutics, Uniformed Services University of the Health Sciences, Bethesda, MD, USA. [80]Pediatric Infectious Diseases and Immunodeficiencies Unit, Vall d'Hebron Barcelona Hospital Campus, Barcelona, Spain. [81]St. Giles Laboratory of Human Genetics of Infectious Diseases, Rockefeller Branch, Rockefeller University, New York, NY, USA; Department of Medical Microbiology, University Medical Center Utrecht, Utrecht, Netherlands. [82]Department of Internal Medicine II, Medical University of Innsbruck, Innsbruck, Austria. [83]Garvan Institute of Medical Research, Darlinghurst, NSW, Australia; St Vincent's Clinical School, Faculty of Medicine, UNSW Sydney, NSW, Australia. [84]Al Jalila Children's Hospital, Dubai, UAE. [85]BC Children's Hospital, University of British Columbia, Vancouver, BC, Canada. [86]Centre for Precision Therapeutics, Genetic and Genomic Medicine Centre, NeuroGen Children Healthcare, Dhaka,

Bangladesh; Holy Family Red Crescent Medical College, Dhaka, Bangladesh. [87]College of Medicine, Mohammed Bin Rashid University of Medicine and Health Sciences, Dubai, UAE; Cellular Intelligence (Ci) Lab, GenomeArc Inc., Toronto, ON, Canada. [88]Department of Neurology, Amsterdam Neuroscience, Amsterdam University Medical Center, University of Amsterdam, Amsterdam, Netherlands. [89]Department of Medicine, Division of Infectious Diseases, McGill University Health Centre, Montréal, QC, Canada; Infectious Disease Susceptibility Program, Research Institute, McGill University Health Centre, Montréal, QC, Canada. [90]Department of Pediatric Pneumology, Immunology and Intensive Care, Charité Universitätsmedizin, Berlin University Hospital Center, Berlin, Germany; Labor Berlin GmbH, Department of Immunology, Berlin, Germany; Berlin Institutes of Health (BIH), Berlin-Brandenburg Center for Regenerative Therapies, Berlin, Germany. [91]Biosciences Institute, University of São Paulo, São Paulo, Brazil. [92]Molecular Biophysics Division, Faculty of Physics, A. Mickiewicz University, Poznań, Poland. [93]Rockefeller University and Howard Hughes Medical Institute, New York, NY, USA; Necker Hospital for Sick Children and INSERM, Paris, France.

**Members of COVID-STORM Clinicians:** Giuseppe Foti[1], Giacomo Bellani[1], Giuseppe Citerio[1], Ernesto Contro[1], Alberto Pesci[2], Maria Grazia Valsecchi[3], Marina Cazzaniga[4]

[1]Department of Emergency, Anesthesia and Intensive Care, School of Medicine and Surgery, University of Milano-Bicocca, San Gerardo Hospital, Monza, Italy. [2]Department of Pneumology, School of Medicine and Surgery, University of Milano-Bicocca, San Gerardo Hospital, Monza, Italy. [3]Center of Bioinformatics and Biostatistics, School of Medicine and Surgery, University of Milano-Bicocca, San Gerardo Hospital, Monza, Italy. [4]Phase I Research Center, School of Medicine and Surgery, University of Milano-Bicocca, San Gerardo Hospital, Monza, Italy.

**Members of COVID Clinicians:** Jorge Abad[1], Giulia Accordino[2], Cristian Achille[3], Sergio Aguilera-Albesa[4], Aina Aguiló-Cucurull[5], Alessandro Aiuti[6], Esra Akyüz Özkan[7], Ilad Alavi Darazam[8], Jonathan Antonio Roblero Albisures[9], Juan C. Aldave[10], Miquel Alfonso Ramos[11], Taj Ali Khan[12], Anna Aliberti[13], Seyed Alireza Nadji[14], Gulsum Alkan[15], Suzan A. AlKhater[16], Jerome Allardet-Servent[17], Luis M. Allende[18], Rebeca Alonso-Arias[19], Mohammed S. Alshahrani[20], Laia Alsina[21], Marie-Alexandra Alyanakian[22], Blanca Amador Borrero[23], Zahir Amoura[24], Arnau Antolí[25], Romain Arrestier[26], Mélodie Aubart[27], Teresa Auguet[28], Iryna Avramenko[29], Gökhan Aytekin[30], Axelle Azot[31], Seiamak Bahram[32], Fanny Bajolle[33], Fausto Baldanti[34], Aurélie Baldolli[35], Maite Ballester[36], Hagit Baris Feldman[37], Benoit Barrou[38], Federica Barzaghi[6], Sabrina Basso[39], Gulsum Iclal Bayhan[40], Alexandre Belot[41], Liliana Bezrodnik[42], Agurtzane Bilbao[43], Geraldine Blanchard-Rohner[44], Ignacio Blanco[45], Adeline Blandinières[46], Daniel Blázquez-Gamero[47], Alexandre Bleibtreu[48], Marketa Bloomfield[49], Mireia Bolivar-Prados[50], Anastasiia Bondarenko[51], Alessandro Borghesi[3], Raphael Borie[52], Elisabeth Botdhlo-Nevers[53], Ahmed A. Bousfiha[54], Aurore Bousquet[55], David Boutolleau[56], Claire Bouvattier[57], Oksana Boyarchuk[58], Juliette Bravais[59], M. Luisa Briones[60], Marie-Eve Brunner[61], Raffaele Bruno[62], Maria Rita P. Bueno[63], Huda Bukhari[64], Jacinta Bustamante[33], Juan José Cáceres Agra[65], Ruggero Capra[66], Raphael Carapito[67], Maria Carrabba[68], Giorgio Casari[6], Carlos Casasnovas[69], Marion Caseris[70], Irene Cassaniti[34], Martin Castelle[71], Francesco Castelli[72], Martín Castillo de Vera[73], Mateus V. Castro[63], Emilie Catherinot[74], Jale Bengi Celik[75], Alessandro Ceschi[76], Martin Chalumeau[77], Bruno Charbit[78], Matthew P. Cheng[79], Père Clavé[50], Bonaventura Clotet[80], Anna Codina[81], Yves Cohen[82], Roger Colobran[83], Cloé Comarmond[84], Alain Combes[85], Patrizia Comoli[39], Angelo G. Corsico[2], Betul Sozeri[86], Taner Coşkuner[86], Aleksandar Cvetkovski[87], Cyril Cyrus[88], David Dalmau[89], François Danion[90], David Ross Darley[91], Vincent Das[92], Nicolas Dauby[93], Stéphane Dauger[94], Paul De Munter[95],

Loic de Pontual[96], Amin Dehban[97], Geoffroy Delplancq[98], Alexandre Demoule[99], Isabelle Desguerre[100], Antonio Di Sabatino[101], Jean-Luc Diehl[102], Stephanie Dobbelaere[103], Elena Domínguez-Garrido[104], Clément Dubost[105], Olov Ekwall[106], Şefika Elmas Bozdemir[107], Marwa H. Elnagdy[108], Melike Emiroglu[15], Akifumi Endo[109], Emine Hafize Erdeniz[110], Selma Erol Aytekin[111], Maria Pilar Etxart Lasa[112], Romain Euvrard[113], Giovanna Fabio[68], Laurence Faivre[114], Antonin Falck[115], Muriel Fartoukh[116], Morgane Faure[117], Miguel Fernandez Arquero[118], Ricard Ferrer[119], Jose Ferreres[120], Carlos Flores[121], Bruno Francois[122], Victoria Fumadó[123], Kitty S. C. Fung[124], Francesca Fusco[125], Alenka Gagro[126], Blanca Garcia Solis[127], Pierre Garçon[345], Pascale Gaussem[128], Zeynep Gayretli[129], Juana Gil-Herrera[130], Laurent Gilardin[131], Audrey Giraud Gatineau[132], Mònica Girona-Alarcón[133], Karen Alejandra Cifuentes Godínez[134], Jean-Christophe Goffard[135], Nacho Gonzales[136], Luis I. Gonzalez-Granado[137], Rafaela González-Montelongo[138], Antoine Guerder[139], Belgin Gülhan[140], Victor Daniel Gumucio[141], Leif Gunnar Hanitsch[142], Jan Gunst[143], Marta Gut[144], Jérôme Hadjadj[145], Filomeen Haerynck[146], Rabih Halwani[147], Lennart Hammarström[148], Selda Hancerli[149], Tetyana Hariyan[150], Nevin Hatipoglu[151], Deniz Heppekcan[152], Elisa Hernandez-Brito[153], Po-ki Ho[154], María Soledad Holanda-Peña[155], Juan P. Horcajada[156], Sami Hraiech[157], Linda Humbert[158], Ivan F. N. Hung[159], Alejandro D. Iglesias[160], Antonio Íñigo-Campos[138], Matthieu Jamme[161], María Jesús Arranz[89], Marie-Thérèse Jimeno[162], Iolanda Jordan[133], Saliha Kanık Yüksek[163], Yalcin Burak Kara[164], Aydın Karahan[165], Adem Karbuz[166], Kadriye Kart Yasar[167], Ozgur Kasapcopur[168], Kenichi Kashimada[169], Sevgi Keles[111], Yasemin Kendir Demirkol[170], Yasutoshi Kido[171], Can Kizil[172], Ahmet Osman Kılıç[173], Adam Klocperk[174], Antonia Koutsoukou[175], Zbigniew J. Król[176], Hatem Ksouri[177], Paul Kuentz[178], Arthur M. C. Kwan[179], Yat Wah M. Kwan[180], Janette S. Y. Kwok[181], Jean-Christophe Lagier[182], David S. Y. Lam[183], Vicky Lampropoulou[184], Fanny Lanternier[185], Yu-Lung Lau[186], Fleur Le Bourgeois[94], Yee-Sin Leo[187], Rafael Leon Lopez[188], Daniel Leung[186], Michael Levin[189], Michael Levy[94], Romain Lévy[33], Zhi Li[78], Daniele Lilleri[34], Edson Jose Adrian Bolanos Lima[190], Agnes Linglart[191], Eduardo López-Collazo[192], José M. Lorenzo-Salazar[138], Céline Louapre[193], Catherine Lubetzki[193], Kwok-Cheung Lung[194], Charles-Edouard Luyt[195], David C. Lye[196], Cinthia Magnone[197], Davood Mansouri[198], Enrico Marchioni[199], Carola Marioli[2], Majid Marjani[200], Laura Marques[201], Jesus Marquez Pereira[202], Andrea Martín-Nalda[203], David Martínez Pueyo[204], Javier Martinez-Picado[205], Iciar Marzana[206], Carmen Mata-Martínez[207], Alexis Mathian[24], Larissa R. B. Matos[63], Gail V. Matthews[208], Julien Mayaux[209], Raquel McLaughlin-Garcia[210], Philippe Meersseman[211], Jean-Louis Mège[212], Armand Mekontso-Dessap[213], Isabelle Melki[115], Federica Meloni[2], Jean-François Meritet[214], Paolo Merlani[215], Özge Metin Akcan[216], Isabelle Meyts[217], Mehdi Mezidi[218], Isabelle Migeotte[219], Maude Millereux[220], Matthieu Million[221], Tristan Mirault[222], Clotilde Mircher[223], Mehdi Mirsaeidi[224], Yoko Mizoguchi[225], Bhavi P. Modi[226], Francesco Mojoli[13], Elsa Moncomble[227], Abián Montesdeoca Melián[228], Antonio Morales Martinez[229], Francisco Morandeira[230], Pierre-Emmanuel Morange[231], Cléemence Mordacq[158], Guillaume Morelle[232], Stéphane J. Mouly[233], Adrián Muñoz-Barrera[138], Cyril Nafati[234], Shintaro Nagashima[235], Yu Nakagama[171], Bénédicte Neven[236], João Farela Neves[237], Lisa F. P. Ng[238], Yuk-Yung Ng[239], hubert Nielly[105], Yeray Novoa Medina[210], Esmeralda Nuñez Cuadros[240], J. Gonzalo Ocejo-Vinyals[241], Keisuke Okamoto[109], Mehdi Oualha[33], Amani Ouedrani[22], Tayfun Özçelik[242], Aslinur Ozkaya-Parlakay[140], Michele Pagani[13], Qiang Pan-Hammarström[148], Maria Papadaki[243], Christophe Parizot[209], Philippe Parola[244], Tiffany Pascreau[245], Stéphane Paul[246], Estela Paz-Artal[247], Sigifredo Pedraza[248], Nancy Carolina González Pellecer[134], Silvia Pellegrini[249], Rebeca Pérez de Diego[127], Xosé Luis Pérez-Fernández[141], Aurélien Philippe[250], Quentin Philippot[116], Adrien Picod[251], Marc Pineton de Chambrun[85], Antonio Piralla[34], Laura Planas-Serra[252], Dominique Ploin[253], Julien Poissy[254], Géraldine Poncelet[70], Garyphallia Poulakou[175], Marie S. Pouletty[255], Persia Pourshahnazari[256], Jia Li Qiu-Chen[257], Paul Quentric[209], Thomas Rambaud[258], Didier Raoult[212], Violette Raoult[259],

Anne-Sophie Rebillat[223], Claire Redin[260], Léa Resmini[261], Pilar Ricart[262], Jean-Christophe Richard[263], Raúl Rigo-Bonnin[264], Nadia rivet[46], Jacques G. Rivière[265], Gemma Rocamora-Blanch[25], Mathieu P. Rodero[266], Carlos Rodrigo[267], Luis Antonio Rodriguez[190], Carlos Rodriguez-Gallego[268], Agustí Rodriguez-Palmero[269], Carolina Soledad Romero[270], Anya Rothenbuhler[271], Damien Roux[272], Nikoletta Rovina[175], Flore Rozenberg[273], Yvon Ruch[90], Montse Ruiz[274], Maria Yolanda Ruiz del Prado[275], Juan Carlos Ruiz-Rodriguez[119], Joan Sabater-Riera[141], Kai Saks[276], Maria Salagianni[184], Oliver Sanchez[277], Adrián Sánchez-Montalvá[278], Silvia Sánchez-Ramón[279], Laire Schidlowski[280], Agatha Schluter[252], Julien Schmidt[281], Matthieu Schmidt[282], Catharina Schuetz[283], Cyril E. Schweitzer[284], Francesco Scolari[285], Anna Sediva[286], Luis Seijo[287], Analia Gisela Seminario[42], Damien Sene[23], Piseth Seng[221], Sevtap Senoglu[167], Mikko Seppänen[288], Alex Serra Llovich[289], Mohammad Shahrooei[97], Anna Shcherbina[290], Virginie Siguret[291], Eleni Siouti[292], David M. Smadja[293], Nikaia Smith[78], Ali Sobh[294], Xavier Solanich[25], Jordi Solé-Violán[295], Catherine Soler[296], Pere Soler-Palacín[297], Betül Sözeri[86], Giulia Maria Stella[2], Yuriy Stepanovskiy[298], Annabelle Stoclin[299], Fabio Taccone[219], Yacine Tandjaoui-Lambiotte[300], Jean-Luc Taupin[301], Simon J. Tavernier[302], Loreto Vidaur Tello[112], Benjamin Terrier[303], Guillaume Thiery[304], Christian Thorball[260], Karolina Thorn[305], Caroline Thumerelle[158], Imran Tipu[306], Martin Tolstrup[307], Gabriele Tomasoni[308], Julie Toubiana[77], Josep Trenado Alvarez[309], Vasiliki Triantafyllia[310], Sophie Trouillet-Assant[311], Jesús Troya[312], Owen T. Y. Tsang[313], Liina Tserel[314], Eugene Y. K. Tso[315], Alessandra Tucci[316], Şadiye Kübra Tüter Öz[15], Matilde Valeria Ursini[125], Takanori Utsumi[225], Yurdagul Uzunhan[317], Pierre Vabres[318], Juan Valencia-Ramos[319], Ana Maria Van Den Rym[127], Isabelle Vandernoot[320], Valentina Velez-Santamaria[321], Silvia Patricia Zuniga Veliz[134], Mateus C. Vidigal[322], Sébastien Viel[253], Cédric Villain[323], Marie E. Vilaire-Meunier[223], Judit Villar-García[324], Audrey Vincent[57], Guillaume Voiriot[326], Alla Volokha[327], Fanny Vuotto[158], Els Wauters[328], Joost Wauters[329], Alan K. L. Wu[330], Tak-Chiu Wu[331], Aysun Yahşi[332], Osman Yesilbas[333], Mehmet Yildiz[168], Barnaby E. Young[187], Ufuk Yükselmiş[334], Mayana Zatz[63], Marco Zecca[39], Valentina Zuccaro[62], Jens Van Praet[335], Bart N. Lambrecht[336], Eva Van Braeckel[336], Cédric Bosteels[336], Levi Hoste[337], Eric Hoste[338], Fré Bauters[336], Jozefien De Clercq[336], Catherine Heijmans[339], Hans Slabbynck[340], Leslie Naesens[341], Benoit Florkin[342], Cécile Boulanger[343], Dimitri Vanderlinden[344]

[1]Germans Trias i Pujol University Hospital and Research Institute, Badalona, Barcelona, Spain. [2]Respiratory Diseases Division, IRCCS Policlinico San Matteo Foundation, University of Pavia, Pavia, Italy. [3]Neonatal Intensive Care Unit, Fondazione IRCCS Policlinico San Matteo, Pavia, Italy. [4]Navarra Health Service Hospital, Pamplona, Spain. [5]Jeffrey Modell Diagnostic and Research Center for Primary Immunodeficiencies, Barcelona, Catalonia, Spain; Immunology Division, Genetics Department, Vall d'Hebron University Hospital (HUVH), Vall d'Hebron Research Institute (VHIR), Vall d'Hebron Barcelona Hospital Campus, Universitat Autònoma de Barcelona (UAB), Barcelona, Catalonia, Spain. [6]Immunohematology Unit, San Raffaele Hospital, Milan, Italy. [7]Ondokuz Mayıs University Medical Faculty Pediatrics, Samsun, Turkey. [8]Department of Infectious Diseases, Loghman Hakim Hospital, Shahid Beheshti University of Medical Sciences, Tehran, Iran. [9]Hospital Regional de Huehuetenango, “Dr. Jorge Vides de Molina,” Huehuetenango, Guatemala. [10]Hospital Nacional Edgardo Rebagliati Martins, Lima, Peru. [11]Parc Sanitari Sant Joan de Déu, Sant Boi de Llobregat Spain. [12]Khyber Medical University, Khyber Pakhtunkhwa, Pakistan. [13]Anesthesia and Intensive Care, Rianimazione I, Fondazione IRCCS Policlinico San Matteo, Pavia, Italy. [14]Virology Research Center, National Institutes of Tuberculosis and Lung Diseases, Shahid Beheshti University of Medical Sciences, Tehran, Iran. [15]Department of Pediatrics, Division of Pediatric Infectious Diseases, Selcuk University Faculty of Medicine, Konya, Turkey. [16]College of Medicine, Imam Abdulrahman Bin Faisal University, Dammam, Saudi

Arabia; Department of Pediatrics, King Fahad Hospital of the University, Al-Khobar, Saudi Arabia. [17]Intensive Care Unit, Hôpital Européen, Marseille, France. [18]Immunology Department, Hospital 12 de Octubre, Research Institute imas12, Complutense University, Madrid, Spain. [19]Immunology Department, Asturias Central University Hospital, Biosanitary Research Institute of the Principality of Asturias (ISPA), Oviedo, Spain. [20]Emergency and Critical Care Medicine Departments, College of Medicine, Imam AbdulRahman Ben Faisal University, Dammam, Saudi Arabia. [21]Clinical Immunology and Primary Immunodeficiencias Unit, Hospital Sant Joan de Déu, Institut de Recerca Sant Joan de Déu, Barcelona, Spain; Universitat de Barcelona, Barcelona, Spain. [22]Department of Biological Immunology, Necker Hospital for Sick Children, AP-HP and INEM, Paris, France. [23]Internal Medicine Department, Hôpital Lariboisière, AP-HP, Paris, France; Université de Paris, Paris, France. [24]Internal Medicine Department, Pitié-Salpétrière Hospital, Paris, France. [25]Department of Internal Medicine, Hospital Universitari de Bellvitge, IDIBELL, Barcelona, Spain. [26]Service de Médecine Intensive Réanimation, Hôpitaux Universitaires Henri Mondor, AP-HP, Créteil, France; Groupe de Recherche Clinique CARMAS, Faculté de Santé de Créteil, Université Paris Est Créteil, Créteil, France. [27]INSERM U1163, University of Paris, Imagine Institute, Paris, France and Pediatric Neurology Department, Necker-Enfants malades Hospital, AP-HP, Paris, France. [28]Hospital U. de Tarragona Joan XXIII. Universitat Rovira i Virgili (URV). IISPV, Tarragona, Spain. [29]Department of Propedeutics of Pediatrics and Medical Genetics, Danylo Halytsky Lviv National Medical University, Lviv, Ukraine. [30]Department of Immunology and Allergy, Konya City Hospital, Konya, Turkey. [31]Private Practice, Paris, France. [32]INSERM U1109, University of Strasbourg, Strasbourg, France. [33]Necker Hospital for Sick Children, AP-HP, Paris, France. [34]Molecular Virology Unit, Microbiology and Virology Department, Fondazione IRCCS Policlinico San Matteo, Pavia, Italy. [35]Department of Infectious Diseases, CHU de Caen, Caen, France. [36]Consorcio Hospital General Universitario, Valencia, Spain. [37]Genetics Institute, Tel Aviv Sourasky Medical Center and Sackler Faculty of Medicine, Tel Aviv University, Tel Aviv, Israel. [38]Department of Urology, Nephrology, Transplantation, APHP-SU, Sorbonne Université, INSERM U 1082, Paris, France. [39]Cell Factory and Pediatric Hematology-Oncology, Fondazione IRCCS Policlinico San Matteo, Pavia, Italy. [40]Yildirim Beyazit University, Faculty of Medicine, Ankara City Hospital, Children's Hospital, Ankara, Turkey. [41]University of Lyon, CIRI, INSERM U1111, National Referee Centre RAISE, Pediatric Rheumatology, HFME, Hospices Civils de Lyon, Lyon, France. [42]Center for Clinical Immunology, CABA, Buenos Aires, Argentina. [43]Cruces University Hospital, Bizkaia, Spain. [44]Paediatric Immunology and Vaccinology Unit, Geneva University Hospitals and Faculty of Medicine, Geneva, Switzerland. [45]University Hospital and Research Institute "Germans Trias i Pujol," Badalona, Spain. [46]Hematology, Georges Pompidou Hospital, AP-HP, Paris, France. [47]Pediatric Infectious Diseases Unit, Instituto de Investigación Hospital 12 de Octubre (imas12), Hospital Universitario 12 de Octubre, Universidad Complutense, Madrid, Spain. [48]Infectious disease Unit, Pitié-Salpêtrière Hospital, AP-AP, Paris, France. [49]Department of Pediatrics, Thomayer's Hospital, first Faculty of Medicine, Charles University, Prague, Czech Republic; Department of Immunology, Motol University Hospital, Second Faculty of Medicine, Charles University, Prague, Czech Republic. [50]Centro de Investigación Biomédica en Red de Enfermedades Hepàticas y Digestivas (Ciberehd), Hospital de Mataró, Consorci Sanitari del Maresme, Mataró, Spain. [51]Shupyk National Healthcare University of Ukraine, Kyiv, Ukraine. [52]Service de Pneumologie, Hopital Bichat, AP-HP, Paris, France. [53]Department of Infectious Diseases, CIC1408, GIMAP CIRI INSERM U1111, University Hospital of Saint-Etienne, Saint-Etienne, France. [54]Clinical Immunology Unit, Pediatric Infectious Disease Department, Faculty of Medicine and Pharmacy, Averroes University Hospital, LICIA Laboratoire d'immunologie clinique, d'inflammation et d'allergie, Hassann Ii University, Casablanca, Morocco. [55]Bégin

Military Hospital, St Mandé, France. [56]Sorbonne Université, INSERM, Institut Pierre Louis d'Epidémiologie et de Santé Publique (iPLESP), AP-HP, Hôpital Pitié Salpêtrière, Service de Virologie, Paris, France. [57]Endocrinology Unit, AP-HP Hôpitaux Universitaires Paris-Sud, Le Kremlin-Bicêtre, France. [58]Department of Children's Diseases and Pediatric Surgery, I. Horbachevsky Ternopil National Medical University, Ternopil, Ukraine. [59]Pneumology Unit, Tenon Hospital, AP-HP, Paris, France. [60]Department of Respiratory Diseases, Hospital Clínico y Universitario de Valencia, Valencia, Spain. [61]Intensive Care Unit, Réseau Hospitalier Neuchâtelois, Neuchâtel, Switzerland. [62]Infectious Diseases Unit, Fondazione IRCCS Policlinico San Matteo, Pavia, Italy. [63]Human Genome and Stem Cell Research Center, University of São Paulo, São Paulo, Brazil. [64]Department of Internal Medicine, College of Medicine, Imam Abdulrahman Bin Faisal University, Dammam, Saudi Arabia. [65]Hospital Insular, Las Palmas de Gran Canaria, Spain. [66]MS Center, Spedali Civili, Brescia, Italy. [67]Laboratoire d'ImmunoRhumatologie Moléculaire, plateforme GENOMAX, INSERM UMR_S 1109, Faculté de Médecine, ITI TRANSPLANTEX NG, Université de Strasbourg, Strasbourg, France. [68]Fondazione IRCCS Ca′ Granda Ospedale Maggiore Policlinico, Milan, Italy. [69]Neuromuscular Unit, Neurology Department, Hospital Universitari de Bellvitge–IDIBELL and CIBERER, Barcelona, Spain. [70]Hopital Robert Debré, Paris, France. [71]Pediatric Immuno-hematology Unit, Necker Enfants Malades Hospital, AP-HP, Paris, France. [72]Department of Infectious and Tropical Diseases, University of Brescia, ASST Spedali Civili di Brescia, Brescia, Italy. [73]Doctoral Health Care Center, Canarian Health System, Las Palmas de Gran Canaria, Spain. [74]Hôpital Foch, Suresnes, France. [75]Selcuk University Faculty of Medicine, Department of Anesthesiology and Reanimation, Intensive Care Medicine Unit, Konya, Turkey. [76]Division of Clinical Pharmacology and Toxicology, Institute of Pharmacological Sciences of Southern Switzerland, Ente Ospedaliero Cantonale and Faculty of Biomedical Sciences, Università della Svizzera italiana, Lugano, Switzerland. [77]Necker Hospital for Sick Children, Paris University, AP-HP, Paris, France. [78]Pasteur Institute, Paris, France. [79]McGill University Health Centre, Montreal, Canada. [80]University Hospital and Research Institute "Germans Trias i Pujol," IrsiCaixa AIDS Research Institute, UVic-UCC, Badalona, Spain. [81]Clinical Biochemistry, Pathology, Paediatric Neurology and Molecular Medicine Departments and Biobank, Institut de Recerca Sant Joan de Déu and CIBERER-ISCIII, Esplugues, Spain. [82]AP-HP, Avicenne Hospital, Intensive Care Unit, Bobigny, France; University Sorbonne Paris Nord, Bobigny, France; INSERM, U942, F-75010, Paris, France. [83]Hospital Universitari Vall d'Hebron, Barcelona, Spain. [84]Pitié-Salpêtrière Hospital, Paris, France. [85]Service de médecine Intensive Réanimation, Groupe Hospitalier Pitié-Salpêtrière, Sorbonne Université, Paris, France. [86]Umraniye Training and Research Hospital, Istanbul, Turkey. [87]Faculty of Medical Sciences at University "Goce Delcev," Shtip, North Macedonia. [88]Department of Biochemistry, College of Medicine, Imam Abdulrahman Bin Faisal University, Dammam, Saudi Arabia. [89]Fundació Docencia i Recerca Mutua Terrassa, Barcelona, Spain. [90]Maladies Infectieuses et Tropicales, Nouvel Hôpital Civil, CHU Strasbourg, Strasbourg, France. [91]UNSW Medicine, St Vincent's Clinical School, Sydney, NSW, Australia; Department of Thoracic Medicine, St Vincent's Hospital Darlinghurst, Sydney, NSW, Australia. [92]Intensive Care Unit, Montreuil Hospital, Montreuil, France. [93]CHU Saint-Pierre, Université Libre de Bruxelles (ULB), Brussels, Belgium. [94]Pediatric Intensive Care Unit, Robert-Debré University Hospital, AP-HP, Paris, France. [95]General Internal Medicine, University Hospitals Leuven, Leuven, Belgium. [96]Hôpital Jean Verdier, AP-HP, Bondy, France. [97]Specialized Immunology Laboratory of Dr. Shahrooei, Sina Medical Complex, Ahvaz, Iran. [98]Centre de génétique humaine, CHU Besançon, Besançon, France. [99]Sorbonne Université médecine and AP-HP Sorbonne université site Pitié-Salpêtrière, Paris, France. [100]Pediatric Neurology Department, Necker-Enfants Malades Hospital, AP-HP, Paris, France. [101]Department of Internal Medicine, Fondazione IRCCS Policlinico San Matteo,

University of Pavia, Pavia, Italy. [102]Intensive Care Unit, Georges Pompidou Hospital, AP-HP, Paris, France. [103]Department of Pneumology, AZ Delta, Roeselare, Belgium. [104]Molecular Diagnostic Unit, Fundación Rioja Salud, Logroño, La Rioja, Spain. [105]Bégin Military Hospital, Saint Mandé, France. [106]Department of Pediatrics, Institute of Clinical Sciences, Sahlgrenska Academy, University of Gothenburg, Gothenburg, Sweden; Department of Rheumatology and Inflammation Research, Institute of Medicine, Sahlgrenska Academy, University of Gothenburg, Gothenburg, Sweden. [107]Bursa City Hospital, Bursa, Turkey. [108]Department of Medical Biochemistry and Molecular Biology, Faculty of Medicine, Mansoura University, Mansoura, Egypt. [109]Tokyo Medical and Dental University, Tokyo, Japan. [110]Ondokuz Mayıs University Faculty of Medicine, Samsun, Turkey. [111]Necmettin Erbakan University, Meram Medical Faculty, Division of Pediatric Allergy and Immunology, Konya, Turkey. [112]Intensive Care Medicine, Donostia University Hospital, Biodonostia Institute of Donostia, CIBER Enfermedades Respiratorias ISCIII, Donostia, Spain. [113]Internal Medicine, University Hospital Edouard Herriot, Hospices Civils de Lyon, Lyon, France. [114]Centre de Génétique, CHU Dijon, Dijon, France. [115]Robert Debré Hospital, Paris, France. [116]AP-HP Tenon Hospital, Paris, France. [117]Sorbonne Universités, UPMC University of Paris, Paris, France. [118]Department of Clinical Immunology, Hospital Clínico San Carlos, Madrid, Spain. [119]Intensive Care Department, Vall d'Hebron University Hospital (HUVH), Vall d'Hebron Barcelona Hospital Campus, Barcelona, Catalonia, Spain; Shock, Organ Dysfunction and Resuscitation Research Group, Vall d'Hebron Research Institute (VHIR), Vall d'Hebron Barcelona Hospital Campus, Barcelona, Catalonia, Spain. [120]Intensive Care Unit, Hospital Clínico y Universitario de Valencia, Valencia, Spain. [121]Genomics Division, Instituto Tecnológico y de Energías Renovables (ITER), Santa Cruz de Tenerife, Spain; CIBER de Enfermedades Respiratorias, Instituto de Salud Carlos III, Madrid, Spain; Research Unit, Hospital Universitario N.S. de Candelaria, Santa Cruz de Tenerife, Spain; Instituto de Tecnologías Biomédicas (ITB), Universidad de La Laguna, San Cristóbal de La Laguna, Spain, Santa Cruz de Tenerife, Spain. [122]CHU Limoges and INSERM CIC 1435 and UMR 1092, Limoges, France. [123]Infectious Diseases Unit, Department of Pediatrics, Hospital Sant Joan de Déu, Barcelona, Spain; Institut de Recerca Sant Joan de Déu, Spain; Universitat de Barcelona (UB), Barcelona, Spain. [124]Department of Pathology, United Christian Hospital, Hong Kong, China. [125]Institute of Genetics and Biophysics "Adriano Buzzati-Traverso," IGB-CNR, Naples, Italy. [126]Department of Pediatrics, Children's Hospital Zagreb, University of Zagreb School of Medicine, Zagreb, Josip Juraj Strossmayer University of Osijek, Medical Faculty Osijek, Osijek, Croatia. [127]Laboratory of Immunogenetics of Human Diseases, IdiPAZ Institute for Health Research, La Paz Hospital, Madrid, Spain. [128]Hematology, AP-HP, Hopital Européen Georges Pompidou and INSERM UMR-S1140, Paris, France. [129]Faculty of Medicine, Department of Pediatrics, Division of Pediatric Infectious Diseases, Karadeniz Technical University, Trabzon, Turkey. [130]Division of Immunology, Hospital General Universitario and Instituto de Investigación Sanitaria "Gregorio Marañón," Madrid, Spain. [131]Bégin Military Hospital, Bégin, France. [132]Aix Marseille Univ, IRD, AP-HM, SSA, VITROME, IHU Méditerranée Infection, Marseille, France, French Armed Forces Center for Epidemiology and Public Health (CESPA), Marseille, France. [133]Pediatric Intensive Care Unit, Hospital Sant Joan de Déu, Barcelona, Spain. [134]Gestion Integral en Salud, Guatemala. [135]Department of Internal Medicine, Hôpital Erasme, Université Libre de Bruxelles, Brussels, Belgium. [136]Immunodeficiencies Unit, Research Institute Hospital, Madrid, Spain. [137]Primary Immunodeficiencies Unit, Pediatrics, University Hospital 12 de Octubre, Madrid, Spain; School of Medicine Complutense University of Madrid, Madrid, Spain. [138]Genomics Division, Instituto Tecnológico y de Energías Renovables (ITER), Santa Cruz de Tenerife, Spain. [139]Assistance Publique Hôpitaux de Paris, Paris, France. [140]Ankara City Hospital, Ankara, Turkey. [141]Department of Intensive Care, Hospital Universitari de Bellvitge, IDIBELL,

Barcelona, Spain. [142]Immunodeficiency Outpatient Clinic, Institute for Medical Immunology, FOCIS Center of Excellence, Charité Universitätsmedizin Berlin, Germany. [143]Surgical Intensive Care Unit, University Hospitals Leuven, Leuven, Belgium. [144]CNAG-CRG, Barcelona Institute of Science and Technology, Barcelona, Spain. [145]Department of Internal Medicine, National Reference Center for Rare Systemic Autoimmune Diseases, AP-HP, APHP-CUP, Hôpital Cochin, Paris, France. [146]Department of Paediatric Immunology and Pulmonology, Center for Primary Immunodeficiency Ghent, Jeffrey Modell Diagnosis and Research Center, PID Research Lab, Ghent University Hospital, Ghent, Belgium. [147]Sharjah Institute of Medical Research, College of Medicine, University of Sharjah, Sharjah, UAE, Sharjah, UAE. [148]Department of Biosciences and Nutrition, SE14183, Huddinge, Karolinska Institutet, Stockholm, Sweden. [149]Department of Pediatrics (Infectious Diseases), Istanbul Faculty of Medicine, Istanbul University, Istanbul, Turkey. [150]I. Horbachevsky Ternopil National Medical University, Ternopil, Ukraine. [151]Pediatric Infectious Diseases Unit, Bakirkoy Dr. Sadi Konuk Training and Research Hospital, University of Health Sciences, Istanbul, Turkey. [152]Health Sciences University, Darıca Farabi Education and Research Hospital, Kocaeli, Turkey. [153]Department of Immunology, Hospital Universitario de Gran Canaria Dr. Negrín, Canarian Health System, Las Palmas de Gran Canaria, Spain. [154]Department of Paediatrics, Queen Elizabeth Hospital, Hong Kong, China. [155]Intensive Care Unit. Marqués de Valdecilla Hospital, Santander, Spain. [156]Hospital del Mar, Institut Hospital del Mar d'Investigacions Mèdiques (IMIM), UAB, UPF, Barcelona, Spain. [157]Intensive Care Unit, APHM, Marseille, France. [158]CHU Lille, Lille, France. [159]Department of Medicine, University of Hong Kong, Hong Kong, China. [160]Department of Pediatrics, Columbia University, New York, NY, USA. [161]Centre hospitalier intercommunal Poissy Saint Germain en Laye, Poissy, France. [162]IHU Méditerranée Infection, Service de l'Information Médicale, Hôpital de la Timone, Marseille, France. [163]Health Science University Ankara City Hospital, Ankara, Turkey. [164]School of Medicine, General Surgery Department Fevzi Çakmak Mah, Marmara University, Istanbul, Turkey. [165]Mersin City Education and Research Hospital, Mersin, Turkey. [166]Division of Pediatric Infectious Diseases, Prof. Dr. Cemil Tascıoglu City Hospital, Istanbul, Turkey. [167]Departments of Infectious Diseases and Clinical Microbiology, Bakirkoy Dr. Sadi Konuk Training and Research Hospital, University of Health Sciences, Istanbul, Turkey. [168]Department of Pediatric Rheumatology, Istanbul University-Cerrahpasa, Istanbul, Turkey. [169]Department of Pediatrics, Tokyo Medical and Dental University, Tokyo, Japan. [170]Health Sciences University, Umraniye Education and Research Hospital, Istanbul, Turkey. [171]Department of Parasitology and Research Center for Infectious Disease Sciences, Graduate School of Medicine, Osaka City University, Osaka, Japan. [172]Pediatric Infectious Diseases Unit of Osman Gazi University Medical School in Eskişehir, Turkey. [173]Meram Medical Faculty, Necmettin Erbakan University, Konya, Turkey. [174]Department of Immunology, Second Faculty of Medicine, Charles University and University Hospital in Motol, Prague, Czech Republic. [175]ICU, First Department of Respiratory Medicine, National and Kapodistrian University of Athens, Medical School, "Sotiria" General Hospital of Chest Diseases, Athens, Greece. [176]Central Clinical Hospital of the Ministry of Interior and Administration, Warsaw, Poland. [177]Clinique des soins intensifs, HFR Fribourg, Fribourg, Switzerland. [178]Oncobiologie Génétique Bioinformatique, PC Bio, CHU Besançon, Besançon, France. [179]Department of Intensive Care, Tuen Mun Hospital, Hong Kong, China. [180]Paediatric Infectious Disease Unit, Hospital Authority Infectious Disease Center, Princess Margaret Hospital, Hong Kong (Special Administrative Region), China. [181]Department of Pathology, Queen Mary Hospital, Hong Kong, China. [182]Aix Marseille Univ, IRD, MEPHI, IHU Méditerranée Infection, Marseille, France. [183]Department of Paediatrics, Tuen Mun Hospital, Hong Kong, China. [184]Biomedical Research Foundation of the Academy of Athens, Athens,

Greece. [185]Necker Hospital, Paris, France. [186]Department of Paediatrics and Adolescent Medicine, University of Hong Kong, Hong Kong, China. [187]National Centre for Infectious Diseases, Singapore, Singapore. [188]Hospital Universitario Reina Sofía, Cordoba, Spain. [189]Imperial College, London, England. [190]Hospital General San Juan de Dios, Ciudad de Guatemala, Guatemala. [191]Endocrinology and Diabetes for Children, AP-HP, Bicêtre Paris-saclay hospital, Le Kremlin-Bicêtre, France. [192]Innate Immunity Group, IdiPAZ Institute for Health Research, La Paz Hospital, Madrid, Spain. [193]Neurology Unit, AP-HP Pitié-Salpêtrière Hospital, Paris University, Paris, France. [194]Department of Medicine, Pamela Youde Nethersole Eastern Hospital, Hong Kong, China. [195]Intensive Care Unit, AP-HP Pitié-Salpêtrière Hospital, Paris University, Paris, France. [196]National Centre for Infectious Diseases, Singapore, Singapore; Tan Tock Seng Hospital, Singapore, Singapore; Yong Loo Lin School of Medicine, Singapore, Singapore; Lee Kong Chian School of Medicine, Singapore, Singapore. [197]Hospital de Niños Dr. Ricardo Gutierrez, Buenos Aires, Argentina. [198]Department of Clinical Immunology and Infectious Diseases, National Research Institute of Tuberculosis and Lung Diseases, Shahid Beheshti University of Medical Sciences, Tehran, Iran. [199]Neurooncology and Neuroinflammation Unit, IRCCS Mondino Foundation, Pavia, Italy. [200]Clinical Tuberculosis and Epidemiology Research Center, National Research Institute of Tuberculosis and Lung Diseases (NRITLD), Shahid Beheshti University of Medical Sciences, Tehran, Iran. [201]Coordenadora da Unidade de Infeciologia e Imunodeficiências do Serviço de Pediatria, Centro Materno-Infantil do Norte, Porto, Portugal. [202]Hospital Sant Joan de Déu and University of Barcelona, Barcelona, Spain. [203]Pediatric Infectious Diseases and Immunodeficiencies Unit, Hospital Universitari Vall d'Hebron, Vall d'Hebron Research Institute, Vall d'Hebron Barcelona Hospital Campus, Universitat Autònoma de Barcelona (UAB), Barcelona, Catalonia, Spain. [204]Hospital Universitari Mutua de Terrassa, Universitat de Barcelona, Barcelona, Spain. [205]IrsiCaixa AIDS Research Institute, ICREA, UVic-UCC, Research Institute "Germans Trias i Pujol," Badalona, Spain. [206]Department of Laboratory, Cruces University Hospital, Barakaldo, Bizkaia, Spain, Bizkaia, Spain. [207]Intensive Care Unit, Hospital General Universitario "Gregorio Marañón," Madrid, Spain. [208]University of New South Wales, Sydney, NSW, Australia. [209]AP-HP Pitié-Salpêtrière Hospital, Paris, France. [210]Department of Pediatrics, Complejo Hospitalario Universitario Insular-Materno Infantil, Canarian Health System, Las Palmas de Gran Canaria, Spain. [211]Medical Intensive Care Unit, University Hospitals Leuven, Leuven, Belgium. [212]Aix-Marseille University, APHM, Marseille, France. [213]Service de Médecine Intensive Réanimation, Hôpitaux Universitaires Henri Mondor, Assistance Publique–Hôpitaux de Paris (AP-HP), Groupe de Recherche Clinique CARMAS, Faculté de Santé de Créteil, Université Paris Est Créteil, France. [214]AP-HP Cohin Hospital, Paris, France. [215]Department of Critical Care Medicine, Ente Ospedaliero Cantonale, Bellinzona, Switzerland. [216]Necmettin Erbakan University, Meram Medical Faculty, Division of Pediatric Infectious Diseases, Konya, Turke4y. [217]Department of Pediatrics, University Hospitals Leuven, Leuven, Belgium; KU Leuven, Department of Microbiology, Immunology and Transplantation; Laboratory for Inborn Errors of Immunity, KU Leuven, Leuven, Belgium. [218]Hospices Civils de Lyon, Hôpital de la Croix-Rousse, Lyon, France. [219]Hôpital Erasme, Brussels, Belgium. [220]Centre hospitalier de gonesse, Gonesse, France. [221]Aix Marseille Univ, IRD, AP-HM, MEPHI, IHU Méditerranée Infection, Marseille, France. [222]Vascular Medicine, Georges Pompidou Hospital, AP-HP, Paris, France. [223]Institut Jérôme Lejeune, Paris, France. [224]Division of Pulmonary and Critical Care, College of Medicine-Jacksonville, University of Florida, Jacksonville, FL, USA. [225]Department of Pediatrics, Hiroshima University Graduate School of Biomedical and Health Sciences, Hiroshima, Japan. [226]BC Children's Hospital Research Institute, University of British Columbia, Vancouver, BC, Canada. [227]Médecine Intensive Réanimation, Hôpitaux Universitaires Henri Mondor, Assistance Publique–Hôpitaux de Paris (AP-HP), Créteil, France. [228]Guanarteme Health Care

Center, Canarian Health System, Las Palmas de Gran Canaria, Spain. [229]Regional University Hospital of Malaga, Malaga, Spain. [230]Department of Immunology, Hospital Universitari de Bellvitge, IDIBELL, Barcelona, Spain. [231]Aix Marseille Univ, INSERM, INRAE, C2VN, Marseille, France. [232]Department of General Paediatrics, Hôpital Bicêtre, AP-HP, University of Paris Saclay, Le Kremlin-Bicêtre, France. [233]INSERM U1144, Université de Paris, DMU INVICTUS, AP-HP.Nord, Département de Médecine Interne, Lariboisière Hospital, Paris, France. [234]CHU de La Timone, Marseille, France. [235]Department of Epidemiology, Infectious Disease Control and Prevention, Graduate School of Biomedical and Health Sciences, Hiroshima University, Hiroshima, Japan. [236]Pediatric Immunology and Rheumatology Department, Necker Hospital, AP-HP, Paris, France. [237]Centro Hospitalar Universitário de Lisboa Central, Lisbon, Portugal. [238]Infectious Disease Horizontal Technology Centre, A*STAR, Singapore, Singapore; Singapore Immunology Network, A*STAR, Singapore. [239]Department of Medicine and Geriatrics, Tuen Mun Hospital, Hong Kong, China. [240]Regional Universitary Hospital of Malaga, Málaga, Spain. [241]Department of Immunology, Hospital Universitario Marqués de Valdecilla, Santander, Spain. [242]Bilkent University, Department of Molecular Biology and Genetics, Ankara, Turkey. [243]BRFAA, Athens, Greece. [244]IHU Méditerranée Infection, Aix Marseille Univ, IRD, AP-HM, SSA, VITROME, IHU Méditerranée Infection, Marseille, France. [245]L'Hôpital Foch, Suresnes, France. [246]Department of Immunology, CIC1408, GIMAP CIRI INSERM U1111, University Hospital of Saint-Etienne, Saint-Etienne, France. [247]Department of Immunology, Hospital Universitario 12 de Octubre, Instituto de Investigación Sanitaria Hospital 12 de Octubre (imas12), Madrid, Spain. [248]Unit of Biochemistry, Instituto Nacional de Ciencias Médicas y Nutrición Salvador Zubirán, Mexico City, Mexico. [249]Diabetes Research Institute, IRCCS San Raffaele Hospital, Milan, Italy. [250]AP-HP Hôpitaux Universitaires Paris-Sud, Le Kremlin-Bicêtre, France. [251]AP-HP, Avicenne Hospital, Intensive Care Unit, Bobigny, France; INSERM UMR-S 942, Cardiovascular Markers in Stress Conditions (MASCOT), University of Paris, Paris, France. [252]Neurometabolic Diseases Laboratory, IDIBELL-Hospital Duran i Reynals, Barcelona; CIBERER U759, ISCiii Madrid, Spain. [253]Hospices Civils de Lyon, Lyon, France. [254]Univ. Lille, INSERM U1285, CHU Lille, Pôle de médecine intensive-réanimation, CNRS, UMR 8576–Unité de Glycobiologie Structurale et Fonctionnelle, Lille, France. [255]Department of General pediatrics, Robert Debre Hospital, Paris, France. [256]University of British Columbia, Vancouver, BC, Canada. [257]Jeffrey Modell Diagnostic and Research Center for Primary Immunodeficiencies, Barcelona, Catalonia, Spain; Diagnostic Immunology Research Group, Vall d'Hebron Research Institute (VHIR), Vall d'Hebron University Hospital (HUVH), Vall d'Hebron Barcelona Hospital Campus, Barcelona, Catalonia, Spain. [258]AP-HP, Avicenne Hospital, Intensive Care Unit, Bobigny, France; University Sorbonne Paris Nord, Bobigny, France. [259]Centre Hospitalier de Saint-Denis, St Denis, France. [260]Precision Medicine Unit, Lausanne University Hospital and University of Lausanne, Lausanne, Switzerland. [261]Paris Cardiovascular Center, PARCC, INSERM, Université de Paris, Paris, France. [262]Germans Trias i Pujol Hospital, Badalona, Spain. [263]Medical Intensive Care Unit, Hopital de la Croix-Rousse, Hospices Civils de Lyon, Lyon, France. [264]Department of Clinical Laboratory, Hospital Universitari de Bellvitge, IDIBELL, Barcelona, Spain. [265]Pediatric Infectious Diseases and Immunodeficiencies Unit, Hospital Universitari Vall d'Hebron, Vall d'Hebron Research Institute, Vall d'Hebron Barcelona Hospital Campus., Barcelona, Spain. [266]Université de Paris, CNRS UMR-8601, Paris, France; Team Chemistry and Biology, Modeling and Immunology for Therapy, CBMIT, Paris, France. [267]Germans Trias i Pujol University Hospital and Research Institute, Badalona, Spain. [268]Department of Immunology, University Hospital of Gran Canaria Dr. Negrín, Canarian Health System, Las Palmas de Gran Canaria, Spain; Department of Clinical Sciences, University Fernando Pessoa Canarias, Las Palmas de Gran Canaria, Spain. [269]Neurometabolic

Diseases Laboratory, Bellvitge Biomedical Research Institute (IDIBELL), 08908 L'Hospitalet de Llobregat, Barcelona, Spain; University Hospital Germans Trias i Pujol, Badalona, Barcelona, Catalonia, Spain. [270]Consorcio Hospital General Universitario, Valencia, Spain. [271]AP-HP Hôpitaux Universitaires Paris-Sud, Paris, France. [272]Intensive Care Unit, Louis-Mourier Hospital, Colombes, France. [273]Virology Unit, Université de Paris, Cohin Hospital, AP-HP, Paris, France. [274]Neurometabolic Diseases Laboratory and CIBERER U759, Barcelona, Spain. [275]Hospital San Pedro, Logroño, Spain. [276]University of Tartu, Institute of Biomedicine and Translational Medicine, Tartu, Estonia. [277]Respiratory Medicine, Georges Pompidou Hospital, AP-HP, Paris, France. [278]Infectious Diseases Department, International Health Program of the Catalan Institute of Health (PROSICS), Vall d'Hebron University Hospital (HUVH), Vall d'Hebron Barcelona Hospital Campus, Universitat Autónoma de Barcelona, Barcelona, Spain. [279]Hospital Clínico San Carlos and IdSSC, Madrid, Spain. [280]Faculdades Pequeno Príncipe, Instituto de Pesquisa Pelé Pequeno Príncipe, Curitiba, Brazil. [281]AP-HP, Avicenne Hospital, Intensive Care Unit, Bobigny, France. [282]Service de Médecine Intensive Réanimation, Institut de Cardiologie, Hopital Pitié-Salpêtrière, Paris, France. [283]Department of Pediatrics, Medizinische Fakultät Carl Gustav Carus, Technische Universität Dresden, Dresden, Germany. [284]CHRU de Nancy, Hôpital d'Enfants, Vandoeuvre, France. [285]Chair of Nephrology, University of Brescia, Brescia, Italy. [286]Department of Immunology, Second Faculty of Medicine, Charles University and Motol University Hospital, Prague, Czech Republic. [287]Clínica Universidad de Navarra and Ciberes, Madrid, Spain. [288]HUS Helsinki University Hospital, Children and Adolescents, Rare Disease Center, and Inflammation Center, Adult Immunodeficiency Unit, Majakka, Helsinki, Finland. [289]Fundació Docencia i Recerca Mutua Terrassa, Terrassa, Spain. [290]D. Rogachev National Medical and Research Center of Pediatric Hematology, Oncology, Immunoogy, Moscow, Russia. [291]Haematology Laboratory, Lariboisière Hospital, University of Paris, Paris, France. [292]Biomedical Research Foundation of the Academy of Athens, Athens, Greece. [293]INSERM U1140, University of Paris, European Georges Pompidou Hospital, Paris, France. [294]Department of Pediatrics, Faculty of Medicine, Mansoura University, Mansoura, Egypt. [295]Intensive Care Medicine, Hospital Universitario de Gran Canaria Dr. Negrín, Canarian Health System, Las Palmas de Gran Canaria, Spain. [296]CHU de Saint Etienne, Saint-Priest-en-Jarez, France. [297]Pediatric Infectious Diseases and Immunodeficiencies Unit, Hospital Universitari Vall d'Hebron, Vall d'Hebron Research Institute, Vall d'Hebron Barcelona Hospital Campus. Universitat Autònoma de Barcelona (UAB), Barcelona, Catalonia, Spain; EU, Barcelona, Spain. [298]Department of Pediatric Infectious Diseases and Pediatric Immunology, Shupyk National Healthcare University of Ukraine, Kyiv, Ukraine. [299]Gustave Roussy Cancer Campus, Villejuif, France. [300]Intensive Care Unit, Avicenne Hospital, AP-HP, Bobigny, France. [301]Laboratory of Immunology and Histocompatibility, Saint-Louis Hospital, Paris University, Paris, France. [302]Center for Inflammation Research, Laboratory of Molecular Signal Transduction in Inflammation, VIB, Ghent, Belgium. [303]Department of Internal Medicine, Université de Paris, INSERM, U970, PARCC, F-75015, Paris, France. [304]Service de médecine intensive réanimation, CHU de Saint-Etienne, France. [305]Department of Rheumatology and Inflammation Research, Institute of Medicine, Sahlgrenska Academy, University of Gothenburg, Gothenburg, Sweden. [306]University of Management and Technology, Lahore, Pakistan. [307]Department of Infectious Diseases, Aarhus University Hospital, Aarhus, Denmark. [308]First Division of Anesthesiology and Critical Care Medicine, University of Brescia, ASST Spedali Civili di Brescia, Brescia, Italy. [309]Intensive Care Department, Hospital Universitari MutuaTerrassa, Universitat Barcelona, Terrassa, Spain. [310]Laboratory of Immunobiology, Center for Clinical, Experimental Surgery and Translational Research, Biomedical Research Foundation of the Academy of Athens, Athens, Greece. [311]International Center of Research in Infectiology, Lyon University, INSERM U1111,

CNRS UMR 5308, ENS, UCBL, Lyon, France; Hospices Civils de Lyon, Lyon Sud Hospital, Pierre-Bénite, France. [312]Infanta Leonor University Hospital, Madrid, Spain. [313]Department of Medicine and Geriatrics, Princess Margaret Hospital, Hong Kong, China. [314]University of Tartu, Institute of Clinical Medicine, Tartu, Estonia. [315]Department of Medicine, United Christian Hospital, Hong Kong, China. [316]Hematology Department, ASST Spedali Civili di Brescia, Brescia, Italy. [317]Pneumologie, Hôpital Avicenne, AP-HP, INSERM U1272, Université Sorbonne Paris Nord, Bobigny, France. [318]Dermatology Unit, Laboratoire GAD, INSERM UMR1231 LNC, Université de Bourgogne, Dijon, France. [319]University Hospital of Burgos, Burgos, Spain. [320]Center of Human Genetics, Hôpital Erasme, Université Libre de Bruxelles, Brussels, Belgium. [321]Bellvitge University Hospital, L'Hospitalet de Llobregat, Barcelona, Spain. [322]University of São Paulo, São Paulo, Brazil. [323]CHU de Caen, Caen, France. [324]Hospital del Mar–IMIM Biomedical Research Institute, Barcelona, Catalonia, Spain. [326]Sorbonne Université, Service de Médecine Intensive Réanimation, Hôpital Tenon, Assistance Publique-Hôpitaux de Paris, Paris, France. [327]Pediatric Infectious Disease and Pediatric Immunology Department, Shupyk National Healthcare University of Ukraine, Kyiv, Ukraine. [328]Department of Pneumology, University Hospitals Leuven, Leuven, Belgium. [329]Laboratory for Clinical Infectious and Inflammatory Disorders, Department of Microbiology, Immunology, and Transplantation, Leuven, Belgium. [330]Department of Clinical Pathology, Pamela Youde Nethersole Eastern Hospital, Hong Kong, China. [331]Department of Medicine, Queen Elizabeth Hospital, Hong Kong, China. [332]Ankara City Hospital, Children's Hospital, Ankara, Turkey. [333]Division of Pediatric Infectious Disease, Department of Pediatrics, Faculty of Medicine, Karadeniz Technical University, Trabzon, Turkey. [334]Health Sciences University, Lütfi Kırdar Kartal Education and Research Hospital, İstanbul, Turkey. [335]Department of Nephrology and Infectiology, AZ Sint-Jan, Bruges, Belgium. [336]Department of Pulmonology, Ghent University Hospital, Belgium. [337]Department of Pediatric Pulmonology and Immunology, Ghent University Hospital, Ghent, Belgium. [338]Department of Intensive Care Unit, Ghent University Hospital, Ghent, Belgium. [339]Department of Pediatric Hemato-oncology, Jolimont Hospital, La louvière, Belgium. [340]Department of Pulmonology, ZNA Middelheim, Antwerp, Belgium. [341]Department of Internal Medicine, Ghent University Hospital, Ghent, Belgium. [342]Department of Pediatric Immuno-hémato-rheumatology, CHR Citadelle, Liége, Belgium. [343]Department of Pediatric Hemato-oncology, UCL Louvain, Brussels, Belgium. [344]Department of Pediatrics, Saint Luc, UCL Louvain, Brussels Belgium. [345]Intensive Care Unit, Grand Hôpital de l'Est Francilien Site de Marne-La-Vallée, Jossigny, France.

**Members of Orchestra Working Group:** Laurent Abel[1], Matilda Berkell[2], Valerio Carelli[3], Alessio Fiorentino[3], Surbi Malhotra[2], Alessandro Mattiaccio[3], Tommaso Pippucci[3], Marco Seri[3], Evelina Tacconelli[4]

[1]Inserm, University Paris cité, Imagine Institute, Paris, France, [2]University of Antwerp, Antwerp, Belgium, [3]University of Bologna, Bologna, 40138, Italy, [4]University of Verona, 37129 Verona, Italy

**Members of French COVID Cohort Study Group:** Laurent Abel[1], Claire Andrejak[2], François Angoulvant[3], Delphine Bachelet[4], Marie Bartoli[5], Romain Basmaci[6], Sylvie Behilill[7], Marine Beluze[8], Dehbia Benkerrou[9], Krishna Bhavsar[4], Lila Bouadma[4], Sabelline Bouchez[10], Maude Bouscambert[11], Minerva Cervantes-Gonzalez[4], Anissa Chair[4], Catherine Chirouze[12], Alexandra Coelho[13], Camille Couffignal[4], Sandrine Couffin-Cadiergues[14], Eric d'Ortenzio[5], Marie-Pierre Debray[4], Lauren Deconinck[4], Dominique Deplanque[15], Diane Descamps[4], Mathilde Desvallée[16], Alpha Diallo[5], Alphonsine Diouf[13], Céline Dorival[9], François Dubos[17],

Xavier Duval[4], Brigitte Elharrar[18], Philippine Eloy[4], Vincent Enouf[7], Hélène Esperou[14], Marina Esposito-Farese[4], Manuel Etienne[19], Eglantine Ferrand Devouge[19], Nathalie Gault[4], Alexandre Gaymard[11], Jade Ghosn[4], Tristan Gigante[20], Morgane Gilg[20], Jérémie Guedj[21], Alexandre Hoctin[13], Isabelle Hoffmann[4], Ikram Houas[14], Jean-Sébastien Hulot[22], Salma Jaafoura[14], Ouifiya Kafif[4], Florentia Kaguelidou[23], Sabrina Kali[4], Antoine Khalil[4], Coralie Khan[16], Cédric Laouénan[4], Samira Laribi[4], Minh Le[4], Quentin Le Hingrat[4], Soizic Le Mestre[5], Hervé Le Nagard[24], François-Xavier Lescure[4], Sophie Letrou[4], Yves Levy[25], Bruno Lina[11], Guillaume Lingas[24], Jean Christophe Lucet[4], Denis Malvy[26], Marina Mambert[13], France Mentré[4], Amina Meziane[9], Hugo Mouquet[7], Jimmy Mullaert[4], Nadège Neant[24], Duc Nguyen[26], Marion Noret[27], Saad Nseir[17], Aurélie Papadopoulos[14], Christelle Paul[5], Nathan Peiffer-Smadja[4], Thomas Perpoint[28], Ventzislava Petrov-Sanchez[5], Gilles Peytavin[4], Huong Pham[4], Olivier Picone[6], Valentine Piquard[4], Oriane Puéchal[29], Christian Rabaud[30], Manuel Rosa-Calatrava[11], Bénédicte Rossignol[20], Patrick Rossignol[30], Carine Roy[4], Marion Schneider[4], Richa Su[4], Coralie Tardivon[4], Marie-Capucine Tellier[4], François Téoulé[9], Olivier Terrier[11], Jean-François Timsit[4], Christelle Tual[31], Sarah Tubiana[4], Sylvie Van Der Werf[7], Noémie Vanel[32], Aurélie Veislinger[31], Benoit Visseaux[4], Aurélie Wiedemann[25], Yazdan Yazdanpanah[4]

[1]INSERM UMR 1163, Paris, France. [2]CHU Amiens, Amiens, France. [3]Hôpital Necker, Paris, France. [4]Hôpital Bichat, Paris, France. [5]ANRS, Paris, France. [6]Hôpital Louis Mourier, Colombes, France. [7]Pasteur Institute, Paris, France. [8]F-CRIN Partners Platform, Paris, France. [9]INSERM UMR 1136, Paris, France. [10]CHU Nantes, France. [11]INSERM UMR 1111, Lyon, France. [12]CHRU Jean Minjoz, Besançon, France. [13]INSERM UMR 1018, Paris, France. [14]INSERM Sponsor, Paris, France. [15]Centre d'Investigation Clinique, INSERM CIC 1403, Centre Hospitalo universitaire de Lille, Lille, France. [16]INSERM UMR 1219, Bordeaux, France. [17]CHU Lille, Lille, France. [18]CHI de Créteil, Créteil, France. [19]CHU Rouen, Rouen, France. [20]F-CRIN INI-CRCT, Nancy, France. [21]Université de Paris, INSERM, IAME, F-75018 Paris, France. [22]Hôpital Européen Georges Pompidou, Paris, France. [23]Hôpital Robert Debré, Paris, France. [24]INSERM UMR 1137, Paris, France. [25]Vaccine Research Institute (VRI), INSERM UMR 955, Créteil, France. [26]CHU Bordeaux, Bordeaux, France. [27]RENARCI, Annecy, France. [28]CHU Lyon, Lyon, France. [29]REACTing, Paris, France. [30]CHU Nancy, Nancy, France. [31]INSERM CIC-1414, Rennes, France. [32]Hôpital la Timone, Marseille, France.

**Members of CoV-Contact Cohort:** Loubna Alavoine[1], Sylvie Behillil[2], Charles Burdet[3], Charlotte Charpentier[4], Aline Dechanet[5], Diane Descamps[6], Xavier Duval[7], Jean-Luc Ecobichon[1], Vincent Enouf[8], Wahiba Frezouls[1], Nadhira Houhou[5], Ouifiya Kafif[5], Jonathan Lehacaut[1], Sophie Letrou[1], Bruno Lina[9], Jean-Christophe Lucet[10], Pauline Manchon[5], Mariama Nouroudine[1], Valentine Piquard[5], Caroline Quintin[1], Michael Thy[11], Sarah Tubiana[1], Sylvie van der Werf[8], Valérie Vignali[1], Benoit Visseaux[10], Yazdan Yazdanpanah[10], Abir Chahine[12], Nawal Waucquier[12], Maria-Claire Migaud[12], Dominique Deplanque[12], Félix Djossou[13], Mayka Mergeay-Fabre[14], Aude Lucarelli[15], Magalie Demar[13], Léa Bruneau[16], Patrick Gérardin[17], Adrien Maillot[16], Christine Payet[18], Bruno Laviolle[19], Fabrice Laine[19], Christophe Paris[19], Mireille Desille-Dugast[19], Julie Fouchard[19], Denis Malvy[20], Duc Nguyen[20], Thierry Pistone[20], Pauline Perreau[20], Valérie Gissot[21], Carole L. E. Goas[21], Samatha Montagne[22], Lucie Richard[23], Catherine Chirouze[24], Kévin Bouiller[24], Maxime Desmarets[25], Alexandre Meunier[26], Marilou Bourgeon[26], Benjamin Lefévre[27], Hélène Jeulin[28], Karine Legrand[29], Sandra Lomazzi[30], Bernard Tardy[31], Amandine Gagneux-Brunon[32], Frédérique Bertholon[33], Elisabeth Botelho-Nevers[32], Kouakam Christelle[34], Leturque Nicolas[34], Layidé Roufai[34], Karine Amat[35], Sandrine Couffin-Cadiergues[34], Hélène Espérou[36], Samia Hendou[34]

[1]Centre d'Investigation Clinique, INSERM CIC 1425, Hôpital Bichat Claude Bernard, AP-HP, Paris, France. [2]Institut Pasteur, Paris, France. [3]Université de Paris, IAME, INSERM U1137,

Paris, France; Hôpital Bichat Claude Bernard, AP-HP, Paris, France. [4]Service de Virologie, Université de Paris, INSERM, IAME, UMR 1137, Hôpital Bichat Claude Bernard, AP-HP, Paris, France. [5]Hôpital Bichat Claude Bernard, AP-HP, Paris, France. [6]IAME INSERM U1140, Hôpital Bichat Claude Bernard, AP-HP, Paris, France. [7]Centre d'Investigation Clinique, INSERM CIC 1425, AP-HP, IAME, Paris University, Paris, France. [8]Institut Pasteur, U3569 CNRS, Université de Paris, Paris, France. [9]Virpath Laboratory, International Center of Research in Infectiology, Lyon University, INSERM U1111, CNRS U5308, ENS, UCBL, Lyon, France. [10]IAME INSERM U1138, Hôpital Bichat Claude Bernard, AP-HP, Paris, France. [11]Center for Clinical Investigation, Assistance Publique-Hôpitaux de Paris, Bichat-Claude Bernard University Hospital, Paris, France. [12]Centre d'Investigation Clinique, INSERM CIC 1403, Centre Hospitalo universitaire de Lille, Lille, France. [13]Service des maladies infectieuses, Centre Hospitalo universitaire de Cayenne, Guyane, France. [14]Centre d'Investigation Clinique, INSERM CIC 1424, Centre Hospitalier de Cayenne, Cayenne, Guyane Française. [15]Service Hôpital de jour Adulte, Centre Hospitalier de Cayenne, Guyane, France. [16]Centre d'Investigation Clinique, INSERM CIC 1410, Centre Hospitalo universitaire de la Réunion, La Réunion, France. [17]Centre d'Investigation Clinique, INSERM CIC 1410, CHU Reunion, Saint-Pierre, Reunion Island. [18]Centre d'Investigation Clinique, INSERM CIC 1410, Centre de Ressources Biologiques, Centre Hospitalo universitaire de la Réunion, La Réunion, France. [19]Centre d'Investigation Clinique, INSERM CIC 1414, Centre Hospitalo universitaire de Rennes, Rennes, France. [20]Service des maladies infectieuses, Centre Hospitalo universitaire de Bordeaux, Bordeaux, France. [21]Centre d'Investigation Clinique, INSERM CIC 1415, CHRU Tours, Tours, France. [22]CRBT, Centre Hospitalo universitaire de Tours, Tours, France. [23]Pole de Biologie Médicale, Centre Hospitalo universitaire de Tours, Tours, France. [24]Service des maladies infectieuses, Centre Hospitalo universitaire de Besançon, Besançon, France. [25]Service des maladies infectieuses, Centre d'investigation clinique, INSERM CIC1431, Centre Hospitalier Universitaire de Besançon, Besançon, France. [26]Centre de Ressources Biologiques–Filière Microbiologique de Besançon, Centre Hospitalier Universitaire, Besançon, France. [27]Université de Lorraine, CHRU-Nancy and APEMAC, Infectious and Tropical Diseases, Nancy, France. [28]Laboratoire de Virologie, CHRU de Nancy Brabois, Vandoeuvre-lès-Nancy, France. [29]INSERM CIC-EC 1433, Centre Hospitalo universitaire de Nancy, Nancy, France. [30]Centre de ressources Biologiques, Centre Hospitalo universitaire de Nancy, Nancy, France. [31]Centre d'Investigation Clinique, INSERM CIC 1408, Centre Hospitalo universitaire de Saint Etienne, Saint Etienne, France. [32]Service des maladies infectieuses, Centre Hospitalo universitaire de Saint Etienne, Saint Etienne, France. [33]Service des maladies infectieuses, CRB[42]-BTK, Centre Hospitalo Universitaire de Saint Etienne, Saint Etienne, France. [34]Pole Recherche Clinique, INSERM, Paris, France. [35]IMEA Fondation Léon M'Ba, Paris, France. [36]INSERM Clinical Research Department, Paris, France.

**Members of the COVIDeF study group:** Serge Bureau[1], Yannick Vacher[1], Anne Gysembergh-Houal[1], Lauren Demerville[1], Abla Chachoua[1], Sebastien Abad[2], Radhiya Abassi[3], Abdelrafie Abdellaoui[3], Abdelkrim Abdelmalek[4], Hendy Abdoul[5], Helene Abergel[6], Fariza Abeud[7], Sophie Abgrall[8], Noemie Abisror[4], Marylise Adechian[9], Nordine Aderdour[9], Hakeem Farid Admane[4], Frederic Adnet[2], Sara Afritt[5], Helene Agostini[10], Claire Aguilar[11], Sophie Agut[12], Tommaso Francesco Aiello[13], Marc Ait Kaci[14], Hafid Ait Oufella[4], Gokula Ajeenthiravasan[15], Virginie Alauzy[3], Fanny Alby-Laurent[11], Lucie Allard[2], Marie-Alexandra Alyanakian[11], Blanca Amador Borrero[7], Sabrina Amam[6], Lucile Amrouche[11], Marc Andronikof[16], Dany Anglicheau[11], Nadia Anguel[9], Djillali Annane[15], Mohammed Aounzou[3], Caroline Aparicio[7], Gladys Aratus[4], Jean-Benoit Arlet[14], Jeremy Arzoine[3], Elisabeth Aslangul[13], Mona Assefi[3], Adeline Aubry[3], Laetitia Audiffred[4], Etienne Audureau[17], Christelle Nathalie Auger[5], Jean-Charles Auregan[8], Celine Awotar[11], Sonia Ayllon Milla[5], Delphine

Azan[5], Laurene Azemar[7], Billal Azzouguen[7], Marwa Bachir Elrufaai[12], Aïda Badsi[7], Prissile Bakouboula[11], Coline Balcerowiak[3], Fanta Balde[12], Elodie Baldivia[17], Eliane-Flore Bangamingo[18], Amandine Baptiste[3], Fanny Baran-Marszak[2], Caroline Barau[17], Nathalie Barget[19], Flore Baronnet[3], Romain Barthelemy[7], Jean-Luc Baudel[4], Camille Baudry[2], Elodie Baudry[9], Laurent Beaugerie[4], Adel Belamri[3], Nicolas Belaube[12], Rhida Belilita[3], Pierre Bellassen[3], Rawan Belmokhtar[2], Isabel Beltran[6], Ruben Benainous[2], Mourad Benallaoua[2], Robert Benamouzig[2], Amélie Benbara[19], Jaouad Benhida[3], Anis Benkhelouf[3], Jihene Benlagha[18], Chahinez Benmostafa[14], Skander Benothmane[18], Miassa Bentifraouine[2], Laurence Berard[4], Quentin Bernier[3], Enora Berti[17], Astrid Bertier[9], Laure Berton[7], Simon Bessis[15], Alexandra Beurton[20], Celine Bianco[4], Clara Bianquis[3], Frank Bidar[3], Philippe Blanche[5], Clarisse Blayau[12], Alexandre Bleibtreu[3], Emmanuelle Blin[12], Coralie Bloch-Queyrat[2], Marie-Christophe Boissier[2], Diane Bollens[4], Marion Bolzoni[4], Rudy pierre Bompard[12], Nicolas Bonnet[2], Justine Bonnouvrier[4], Shirmonecrystal Botha[3], Wissam Boucenna[4], Fatiha Bouchama[3], Olivier Bouchaud[2], Hanane Bouchghoul[9], Taoueslylia Boudjebla[12], Noel Boudjema[17], Catherine Bouffard[6], Adrien Bougle[3], Meriem Bouguerra[3], Leila Bouras[7], Agnes Bourcier[3], Anne Bourgarit Durand[19], Anne Bourrier[4], Fabrice Bouscarat[6], Diane Bouvry[2], Nesrine Bouziri[3], Ons Bouzrara[3], Sarah Bribier[9], Delphine Brugier[3], Melanie Brunel[11], Eida Bui[4], Anne Buisson[21], Iryna Bukreyeva[9], Côme Bureau[20], Jacques Cadranel[12], Johann Cailhol[2], Ruxandra Calin[12], Clara Campos Vega[11], Pauline Canavaggio[3], Marta Cancella[3], Delphine Cantin[22], Albert Cao[3], Lionel Carbillon[19], Nicolas Carlier[5], Clementine Cassard[3], Guylaine Castor[7], Marion Cauchy[7], Olivier Cha[4], Benjamin Chaigne[5], Salima Challal[2], Karine Champion[7], Patrick Chariot[19], Julie Chas[12], Simon Chauveau[2], Anthony Chauvin[7], Clement Chauvin[18], Nathalie Chavarot[11], Kamélia Chebbout[3], Mustapha Cherai[3], Ilaria Cherubini[3], Amelie Chevalier[5], Thibault Chiarabini[4], Thierry Chinet[10], Richard Chocron[14], Pascaline Choinier[12], Juliette Chommeloux[3], Christophe Choquet[6], Laure Choupeaux[11], Benjamin Chousterman[7], Dragosmarius Ciocan[8], Ada Clarke[5], Gaëlle Clavere[23], Florian Clavier[3], Karine Clement[3], Sebastien Clerc[14], Yves Cohen[2], Fleur Cohen[3], Adrien Cohen[23], Audrey Coilly[24], Hester Colboc[25], Pauline Colin[3], Magalie Collet[7], Chloé Comarmond[7], Emeline Combacon[5], Alain Combes[3], Celine Comparon[2], Jean-Michel Constantin[3], Hugues Cordel[2], Anne-Gael Cordier[9], Adrien Costantini[10], Nathalie Costedoat Chalumeau[5], Camille Couffignal[6], Doriane Coupeau[4], Alain Creange[17], Yannie Cuvillier Lamarre[22], Charlène Da Silveira[6], Sandrine Dautheville Guibal El Kayani[12], Nathalie De Castro[18], Yann De Rycke[3], Lucie Del Pozo[19], Quentin Delannoy[3], Mathieu Delay[12], Robin Deleris[3], Juliette Delforge[13], Laëtitia Delphine[3], Noemie Demare[2], Sophie Demeret[3], Alexandre Demoule[3], Aurore Deniau[2], François Depret[18], Sophie Derolez[2], Ouda Derradji[9], Nawal Derridj[10], Vincent Descamps[6], Lydia Deschamps[6], Celine Desconclois[8], Cyrielle Desnos[3], Karine Desongins[18], Robin Dhote[2], Benjamin Diallo[12], Morgane Didier[2], Myriam Diemer[7], Stephane Diez[9], Juliette Djadi-Prat[14], Fatima-Zohra Djamouri Monnory[12], Siham Djebara[3], Naoual Djebra[2], Minette Djietcheu[18], Hadjer Djillali[4], Nouara Djouadi[4], Severine Donneger[19], Catarina Dos Santos[5], Nathalie Dournon[2], Martin Dres[20], Laura Droctove[3], Marie Drogrey[3], Margot Dropy[3], Elodie Drouet[4], Valérie Dubosq[12], Evelyne Dubreucq[12], Estelle Dubus[7], Boris Duchemann[2], Thibault Duchenoy[5], Emmanuel Dudoignon[18], Romain Dufau[19], Florence Dumas[5], Clara Duran[10], Emmanuelle Duron[24], Antoine Durrbach[17], Claudine Duvivier[11], Nathan Ebstein[2], Jihane El Khalifa[6], Alexandre Elabbadi[12], Caroline Elie[11], Gabriel Ernotte[3], Anne Esling[11], Martin Etienne[9], Xavier Eyer[7], Muriel sarah Fartoukh[12], Takoua Fayali[3], Marion Fermaut[19], Arianna Fiorentino[4], Souha Fliss[2], Marie-Céline Fournier[7], Benjamin Fournier[11], Hélène Francois[12], Olivia Freynet[2], Yvann Frigout[14], Isaure Fromont[7], Axelle Fuentes[6], Thomas Furet[3], Joris Galand[7], Marc Garnier[4],

Agnes Gaubert[3], Stéphane Gaudry[2], Samuel Gaugain[7], Damien Gauthier[3], Maxime Gautier[7], Sophie Georgin-Lavialle[12], Daniela Geromin[14], Mohamed Ghalayini[2], Bijan Ghaleh[17], Myriam Ghezal[21], Aude Gibelin[12], Linda Gimeno[3], Benoit Girard[5], Bénédicte Giroux Leprieur[2], Doryan Gomes[18], Elisabete Gomes-Pires[11], Guy Gorochov[3], Anne Gouge[18], Amel Gouja[17], Helene Goulet[12], Sylvain Goupil[11], Jeanne Goupil De Bouille[2], Julien Gras[7], Segolene Greffe[10], Lamiae Grimaldi[9], Paul Guedeney[3], Bertrand Guidet[4], Matthias Guillo[18], Mariechristelle Gulczynski[26], Tassadit Hadjam[7], Didier Haguenauer[13], Soumeya Hammal[3], Nadjib Hammoudi[3], Olivier Hanon[23], Anarole Harrois[9], Pierre Hausfater[3], Coraline Hautem[14], Guillaume Hekimian[3], Nicholas Heming[15], Olivier Hermine[11], Sylvie Ho[3], Marie Houllier[9], Benjamin Huot[7], Tessa Huscenot[7], Wafa Ibn Saied[12], Ghilas Ikherbane[3], Meriem Imarazene[11], Patrick Ingiliz[4], Lina Iratni[17], Stephane Jaureguiberry[9], Jean-Francois Jean-Marc[10], Deleena Jeyarajasingham[18], Pauline Jouany[14], Veronique Jouis[7], Clement Jourdaine[7], Ouifiya Kafif[6], Rim Kallala[24], Sandrine Katsahian[14], Lilit Kelesyan[27], Vixra Keo[3], Flora Ketz[21], Warda Khamis[2], Enfel Khelili[3], Mehdi Khellaf[17], Christy Gaëlla Kotokpo Youkou[10], Ilias Kounis[24], Gaelle Kpalma[3], Jessica Krause[4], Vincent Labbe[12], Karine Lacombe[4], Jean-Marc Lacorte[3], Anne Gaelle Lafont[4], Emmanuel Lafont[11], Lynda Lagha[27], Lionel Lamhaut[11], Aymeric Lancelot[3], Cecilia Landman[4], Fanny Lanternier[11], Cecile Larcheveque[3], Caroline Lascoux Combe[18], Ludovic Lassel[12], Benjamin Laverdant[12], Christophe Lavergne[18], Jean-Rémi Lavillegrand[4], Pompilia Lazureanu[7], Loïc Le Guennec[3], Lamia Leberre[4], Claire Leblanc[19], Marion Leboyer[28], Francois Lecomte[5], Marine Lecorre[3], Romain Leenhardt[4], Marylou Lefebvre[4], Bénédicte Lefebvre[4], Paul Legendre[5], Anne Leger[3], Laurence Legros[24], Justyna Legrosse[3], Sébastien Lehuunghia[5], Julien Lemarec[3], Jeremie Leporrier-Ext[11], Manon Lesein[5], Hubert Lesur[24], Vincent Levy[2], Albert Levy[14], Edwige Lopes[7], Amanda Lopes[7], Vanessa Lopez[11], Julien Lopinto[12], Olivier Lortholary[11], Badr Louadah[7], Bénédicte Loze[18], Marie-Laure Lucas[22], Axelle Lucasamichi[8], Liem Binh Luong[5], Arouna Magazimama-Ext[7], David Maingret[7], Lakhdar Mameri[18], Philippe Manivet[7], Cylia Mansouri[4], Estelle Marcault[6], Jonathan Marey[5], Nathalie Marin[5], Clémence Marois[3], Olivier Martin[2], Lou Martineau[3], Cannelle Martinez-Lopez[15], Pierre Martyniuck[4], Pauline Mary De Farcy[29], Nessrine Marzouk[12], Rafik Masmoudi[14], Alexandre Mebazaa[7], Frédéric Mechai[2], Fabio Mecozzi[11], Chamseddine Mediouni[10], Bruno Megarbane[7], Mohamed Meghadecha[22], Élodie Mejean[12], Arsene Mekinian[4], Nour Mekki Abdelhadi[6], Rania Mekni[3], Thinhinan Sabrina Meliti[3], Breno Melo Lima[18], Paris Meng[12], Soraya Merbah[3], Fadhila Messani[2], Yasmine Messaoudi[3], Baboo-Irwinsingh Mewasing[12], Lydia Meziane[3], Carole Michelot-Burger[11], Françoise Mignot[18], Fadi Hillary Minka[7], Makoto Miyara[3], Pierre Moine[15], Jean-Michel Molina[18], Anaïs Montegnies-Boulet[5], Alexandra Monti[21], Claire Montlahuc[18], Anne-Lise Montout[3], Alexandre Moores[5], Caroline Morbieu[5], Helene Mortelette[14], Stéphane Mouly[7], Rosita Muzaffar[18], Cherifa Iness Nacerddine[3], Marine Nadal[12], Hajer Nadif[3], Kladoum Nassarmadji[7], Pierre Natella[17], Sandrine Ndingamondze[3], Stefan Neraal[5], Caroline Nguyen[6], Bao N'Guyen[3], Isabelle Nion Larmurier[4], Luc Nlomenyengue[14], Nicolas Noel[9], Hilario Nunes[2], Edris Omar[3], Zineb Ouazene[4], Elise Ouedraogo[2], Wassila Ouelaa[3], Anissa Oukhedouma[3], Yasmina Ould Amara[3], Herve Oya[3], Johanna Oziel[2], Thomas Padilla[3], Elena Paillaud[26], Solenne Paiva[3], Beatrice Parfait[5], Perrine Parize[11], Christophe Parizot[3], Antoine Parrot[12], Arthur Pavot[9], Laetitia Peaudecerf[5], Frédéric Pene[5], Marion Pepin[10], Julie Pernet[3], Claire Pernin[7], Mylène Petit[2], Olivier Peyrony[18], Marie-Pierre Pietri[22], Olivia Pietri[4], Marc Pineton De Chambrun[3], Michelle Pinson[13], Claire Pintado[18], Valentine Piquard[6], Christine Pires[3], Benjamin Planquette[14], Sandrine Poirier[8], Anne-Laure Pomel[8], Stéphanie Pons[3], Diane Ponscarme[18], Annegaelle Pourcelot[9], Valérie Pourcher[3], Anne Pouvaret[11], Florian Prever[4], Miresta Previlon[18], Margot Prevost[3], Marie-Julie Provoost[7], Cyril Quemeneur[3], Cédric Rafat[12],

Agathe Rami[7], Brigitte Ranque[14], Maurice Raphael[9], Jean Herle Raphalen[11], Anna Rastoin[7], Mathieu Raux[3], Amani Rebai[2], Michael Reby[25], Alexis Regent[5], Asma Regrag[14], Matthieu Resche-Rigon[18], Quentin Ressaire[18], Christian Richard[9], Mariecaroline Richard[3], Maxence Robert[3], Benjamin Rohaut[3], Camille Rolland-Debord[12], Jacques Ropers[3], Anne-Marie Roque-Afonso[24], Charlotte Rosso[30], Mélanie Rousseaux[4], Nabila Rousseaux[3], Swasti Roux[26], Lorène Roux[4], Claire Rouzaud[11], Antoine Rozes[3], Emma Rubenstein[7], Jean-Marc Sabate[2], Sheila Sabet[12], Sophie-Caroline Sacleux[24], Nathalie Saidenberg Kermanach[2], Faouzi Saliba[24], Dominique Salmon[22], Laurent Savale[31], Guillaume Savary[3], Rebecca Sberro[11], Anne Scemla[11], Frederic Schlemmer[17], Mathieu Schwartz[7], Saïd Sedfi[3], Samia Sefir-Kribel[5], Philippe Seksik[4], Pierre Sellier[7], Agathe Selves[3], Nicole Sembach[14], Luca Semerano[2], Marie-Victoire Senat[9], Damien Sene[7], Alexandra Serris[11], Lucile Sese[2], Naima Sghiouar[15], Johanna Sigaux[2], Martin Siguier[12], Johanne Silvain[3], Noémie Simon[3], Tabassome Simon[4], Lina Innes Skandri[2], Miassa Slimani[2], Aurélie Snauwaert[6], Harry Sokol[4], Heithem Soliman[4], Nisrine Soltani[9], Benjamin Soyer[7], Gabriel Steg[6], Lydia Suarez[7], Tali-Anne Szwebel[5], Kossi Taffame[3], Yacine Tandjaoui-Lambiotte[2], Claire Tantet[2], Mariagrazia Tateo[18], Igor Theodose[18], Pierre clement Thiebaud[4], Caroline Thomas[4], Kelly Tiercelet[18], Julie Tisserand[9], Carole Tomczak[18], Krystel Torelino[3], Fatima Touam-Ext[11], Lilia Toumi[11], Gustave Toury[14], Mireille Toy-Miou[3], Olivia Tran Dinh Thanh Lien[7], Alexy Trandinh[6], Jean-Marc Treluyer[5], Baptiste Trinque[7], Jennifer Truchot[5], Florence Tubach[3], Sarah Tubiana[6], Simone Tunesi[19], Matthieu Turpin[12], Agathe Turpin[3], Tomas Urbina[4], Rafael Usubillaga Narvaez[22], Yurdagul Uzunhan[2], Prabakar Vaittinadaayar[27], Arnaud Valent[18], Maelle Valentian[12], Nadia Valin[4], Hélène Vallet[4], Marina Vaz[3], Miguel-Alejandro Vazquezibarra[7], Benoit Vedie[14], Laetitia Velly[3], Celine Verstuyft[9], Cedric Viallette[3], Eric Vicaut[7], Dorothee Vignes[8], Damien Vimpere[11], Myriam Virlouvet[9], Guillaume Voiriot[12], Lena Voisot[21], Emmanuel Weiss[27], Nicolas Weiss[3], Anaïs Winchenne[2], Youri Yordanov[4], Lara Zafrani[18], Mohamad Zaidan[9], Wissem Zaidi[4], Cathia Zak[12], Aida Zarhrate-Ghoul[3], Ouassila Zatout[6], Suzanne Zeino[9], Michel Zeitouni[3], Naïma Zemirli[3], Lorene Zerah[3], Ounsa Zia[3], Marianne Ziol[19], Oceane Zolario[4], Julien Zuber[11]

[1]DRCI-APHP, Paris, France, [2]Hôpital Avicenne, Bobigny, France, [3]Hôpital Pitié-Salpêtrière, Paris, France, [4]Hôpital Saint-Antoine, Paris, France, [5]Hôpital Cochin, Paris, France, [6]Hôpital Bichat, Paris, France, [7]Hôpital Lariboisière, Paris, France, [8]Hôpital Antoine Béclère, Clamart, France, [9]Hôpital Kremlin Bicêtre, Le Kremlin-Bicêtre, France, [10]Hôpital Ambroise-Paré, Boulogne Billancourt, France, [11]Hopital Necker Enfants malades, Paris, France, [12]Hôpital Tenon, Paris, France, [13]Hôpital Louis Mourier, Colombes, France, [14]Hôpital Européen Georges Pompidou, Paris, France, [15]Hôpital Raymond Poincaré, Garches, France, [16]Hôpital Antoine Béclère, Calmart, France, [17]Hôpital Henri Mondor, Créteil, France, [18]Hôpital Saint Louis, Paris, France, [19]Hôpital Jean Verdier, Bondy, France, [20]Université Paris-Sorbonne, Hôpital Pitié-Salpêtrière, INSERM, Paris, France, [21]Hôpital Charles Foix, Ivry-sur-Seine, France, [22]Hôpital Hôtel Dieu, Paris, France, [23]Hôpital Broca, Paris, France, [24]Hôpital Paul-Brousse, Villejuif, France, [25]Hôpital Rothschild, Paris, France, [26]Hôpital Corentin Celton, Issy-les-Moulineaux, France, [27]Hôpital Beaujon, Clichy, France, [28]Hôpital Albert Chenevier, Créteil, France, [29]Hôpital Sainte-Périne, Paris, France, [30]Université Paris-Sorbonne, Hôpital Pitié-Salpêtrière, INSERM, CNRS, Paris, France, [31]Université Paris-Saclay, Hôpital Kremlin Bicêtre, INSERM, Le Kremlin-Bicêtre, France

**Members of Amsterdam UMC Covid-19 Biobank:** Michiel van Agtmael[2], Anne Geke Algera[1], Brent Appelman[2], Frank van Baarle[1], Diane Bax[3], Martijn Beudel[4], Harm Jan Bogaard[5], Marije Bomers[2], Peter Bonta[5], Lieuwe Bos[1], Michela Botta[1], Justin de Brabander[2], Godelieve de Bree[2], Sanne de Bruin[1], David T. P. Buis[1], Marianna Bugiani[5], Esther Bulle[1], Osoul Chouchane[2] Alex Cloherty[3], Mirjam Dijkstra[12], Dave A. Dongelmans[1], Romein W. G. Dujardin[1], Paul Elbers[1], Lucas Fleuren[1], Suzanne Geerlings[2] Theo Geijtenbeek[3], Armand Girbes[1], Bram Goorhuis[2], Martin P. Grobusch[2], Florianne Hafkamp[3], Laura Hagens[1], Jorg Hamann[7], Vanessa Harris[2], Robert Hemke[8], Sabine M. Hermans[2] Leo Heunks[1], Markus Hollmann[6], Janneke Horn[1], Joppe W. Hovius[2], Menno D. de Jong[9], Rutger Koning[4], Endry H. T. Lim[1], Niels van Mourik[1], Jeaninne Nellen[2], Esther J. Nossent[5], Frederique Paulus[1], Edgar Peters[2], Dan A. I. Pina-Fuentes[4], Tom van der Poll[2], Bennedikt Preckel[6], Jan M. Prins[2], Jorinde Raasveld[1], Tom Reijnders[2], Maurits C. F. J. de Rotte[12], Michiel Schinkel[2], Marcus J. Schultz[1], Femke A. P. Schrauwen[12], Alex Schuurmans[10], Jaap Schuurmans[1], Kim Sigaloff[1], Marleen A. Slim[1,2], Patrick Smeele[5], Marry Smit[1], Cornelis S. Stijnis[2], Willemke Stilma[1], Charlotte Teunissen[11], Patrick Thoral[1], Anissa M. Tsonas[1], Pieter R. Tuinman[2], Marc van der Valk[2], Denise Veelo[6], Carolien Volleman[1], Heder de Vries[1], Lonneke A. Vught[1,2], Michèle van Vugt[2], Dorien Wouters[12], A. H. (Koos) Zwinderman[13], Matthijs C. Brouwer[4], W. Joost Wiersinga[2], Alexander P. J. Vlaar[1], Diederik van de Beek[4]

[1]Department of Intensive Care, Amsterdam UMC, Amsterdam, Netherlands. [2]Department of Infectious Diseases, Amsterdam UMC, Amsterdam, Netherlands. [3]Experimental Immunology, Amsterdam UMC, Amsterdam, Netherlands. [4]Department of Neurology, Amsterdam UMC, Amsterdam Neuroscience, Amsterdam, Netherlands. [5]Department of Pulmonology, Amsterdam UMC, Amsterdam, Netherlands. [6]Department of Anesthesiology, Amsterdam UMC, Amsterdam, Netherlands. [7]Amsterdam UMC Biobank Core Facility, Amsterdam UMC, Amsterdam, Netherlands. [8]Department of Radiology, Amsterdam UMC, Amsterdam, Netherlands. [9]Department of Medical Microbiology, Amsterdam UMC, Amsterdam, Netherlands. [10]Department of Internal Medicine, Amsterdam UMC, Amsterdam, Netherlands. [11]Neurochemical Laboratory, Amsterdam UMC, Amsterdam, Netherlands. [12]Department of Clinical Chemistry, Amsterdam UMC, Amsterdam, Netherlands. [13]Department of Clinical Epidemiology, Biostatistics and Bioinformatics, Amsterdam UMC, Amsterdam, Netherlands.

**Members of NIAID-USUHS COVID Study Group:** Miranda F. Tompkins[1], Camille Alba[1], Andrew L. Snow[2], Daniel N. Hupalo[1], John Rosenberger[1], Gauthaman Sukumar[1], Matthew D. Wilkerson[1], Xijun Zhang[1], Justin Lack[3], Andrew J. Oler[4], Kerry Dobbs[5], Ottavia M. Delmonte[5], Jeffrey J. Danielson[5], Andrea Biondi[6], Laura Rachele Bettini[6], Mariella D'Angio'[6], Ilaria Beretta[7], Luisa Imberti[8], Alessandra Sottini[8], Virginia Quaresima[8], Eugenia Quiros-Roldan[9], Camillo Rossi[10]

[1]American Genome Center, Uniformed Services University of the Health Sciences, Bethesda, MD, USA; Henry M. Jackson Foundation for the Advancement of Military Medicine, Bethesda, MD, USA. [2]Department of Pharmacology and Molecular Therapeutics, Uniformed Services University of the Health Sciences, Bethesda, MD, USA. [3]NIAID Collaborative Bioinformatics Resource, Frederick National Laboratory for Cancer Research, Leidos Biomedical Research Inc., Frederick, MD, USA. [4]Bioinformatics and Computational Biosciences Branch, Office of Cyber Infrastructure and Computational Biology, NIAID, NIH, Bethesda, MD, USA. [5]Laboratory of Clinical Immunology and Microbiology, Division of Intramural Research, NIAID, NIH, Bethesda, MD, USA. [6]Pediatric Departement and Centro Tettamanti-European Reference Network PaedCan, EuroBloodNet, MetabERN-University of Milano-Bicocca-Fondazione MBBM-Ospedale, San Gerardo, Monza, Italy. [7]Department of Infectious Diseases,

University of Milano-Bicocca, San Gerardo Hospital, Monza, Italy. [8]CREA Laboratory, Diagnostic Department, ASST Spedali Civili di Brescia, Brescia, Italy. [9]Department of Infectious and Tropical Diseases, University of Brescia and ASST Spedali Civili di Brescia, Brescia, Italy. [10]Chief Medical Officer, ASST Spedali Civili di Brescia, Brescia, Italy.
